# Effectiveness of Implementation Interventions in Musculoskeletal Healthcare: A Systematic Review

**DOI:** 10.1101/2023.11.29.23299209

**Authors:** Peter Bech Hansen, Mikkel Bahnsen, Mikkel Sloth Nørgaard, Jette Frost Jepsen, Michael Skovdal Rathleff, Kristian Damgaard Lyng

## Abstract

**Background:** Implementing new knowledge into clinical practice is a challenge, but nonetheless crucial to improve our healthcare system related to the management of musculoskeletal pain. This systematic review aimed to assess the effectiveness of implementation interventions within musculoskeletal healthcare.

**Methods:** We searched Medline, Embase, Cochrane Central Register of Controlled Trials, and Scopus. Any type of randomised controlled trials investigating implementation strategies or interventions in relation to musculoskeletal pain conditions were included. Risk of bias were assessed using the Cochrane Risk of Bias 2 tool. Data analysis was done using frameworks from Powell et al. 2015, and Waltz et al. 2015 and outcomes were identified by Thompson et al. 2022 or self-made outcome domains were established.

**Results:** The literature search yielded 14,265 original studies, of which 38 studies from 31 trials, with 13,203 participating healthcare professionals and 30,320 participating patients were included in the final synthesis. Nineteen studies had a high risk of bias, sixteen had a moderate risk of bias, and three had a low risk of bias. Twenty distinct implementation interventions were identified. A significant heterogeneity in the utilised outcome measurements was observed, thereby rendering a meta-analysis infeasible; consequently, all outcomes were classified into six outcome domains for healthcare professionals, seven for patients and one for cost-effectiveness.

**Conclusions:** Our findings suggest that some implementation interventions may have a tendency towards a statistically significant positive effect in favour of the intervention group on the outcome domain “Adherence to the implemented interventions” for healthcare professionals in the included studies. The remaining outcome domains yielded varying results; therefore, these findings should be interpreted with caution. Future high-quality trials with clear reporting and rationale of implementation strategies and interventions utilising standardised nomenclature are needed to further advance our understanding of this area.

**Trial registration:** Open Science Framework, DOI: <u>10.17605/OSF.IO/SRMP2</u>

## Background

Musculoskeletal pain conditions are a significant burden on societies and healthcare systems worldwide (1,2). In 2019, musculoskeletal conditions affected 1.71 billion people worldwide, and the prevalence is expected to rise owing to aging populations and changing lifestyles (1,3). In addition, the economic and societal impact is severe due to lost productivity, including sick days, lost labour, early retirement and increased healthcare utilisation, and healthcare systems globally spend a significant proportion of their assigned resources on treating musculoskeletal conditions (4–6). Currently, most healthcare services in the western world face a substantial challenge: unnecessary testing and treatment seem to be abundant and lead to wasted economic resources for both patients and the healthcare system (7–12). As patients with musculoskeletal conditions often seek care from healthcare professionals (HCPs), most countries’ health authorities have issued a series of evidence-based guidelines of best practice for the most common conditions; however, adherence to these guidelines seems to be poor (13). In addition, several global and local initiatives have been launched in an effort to reduce unnecessary testing and treatment (14–17). The literature acknowledges the difficulty in implementing evidence-based guidelines (18). Some studies have demonstrated a significant delay between the development of new knowledge and its uptake in clinical practice (19,20). Therefore, the study of implementation science is crucial for developing effective and efficient implementation strategies to ensure that HCPs across various sectors incorporate this knowledge into musculoskeletal healthcare. An implementation strategy consists of a bundle of two or more implementation interventions, which are defined as “a method or technique designed to enhance adoption of a clinical intervention” (21). Several models and frameworks for implementation have been suggested in the literature that can guide the selection of the most appropriate implementation strategy depending on the task at hand (22,23). However, it remains unclear which implementation strategies and interventions are the most effective in various musculoskeletal healthcare settings. Therefore, the aim of this systematic review was to assess which implementation interventions are effective within musculoskeletal healthcare.

## Methods

### Protocol and Registration

This study protocol were preregistered in Open Science Framework (DOI: <u>10.17605/OSF.IO/SRMP2</u>). The reporting of the study followed the Preferred Reporting Items for Systematic Reviews and Meta-Analyses (PRISMA) guidelines See the completed PRISMA checklist in ***Additional file 1***.

### Deviation from Protocol and Preregistration

Several deviations from the protocol occurred due to various reasons. Firstly, our pre-registration stated that the extracted outcomes would be divided into primary- and secondary outcomes, however, for the sake of simplicity, we categorised outcomes into HCP-related, patient-related, and economic-related outcomes and only extracted primary outcomes for each included study. Secondly, we stated that we intended to have two authors extract data independently, but to ensure feasibility, timeliness, and rigour, one author extracted data which were validated by another author.

### Study Inclusion and Exclusion Criteria

Only randomised control trials (RCTs) were included for this study. Studies were included if they were randomised by individuals, clusters or used a stepped-wedge design. Furthermore, all studies needed to be published in English, include at least one implementation strategy or intervention in relation to musculoskeletal healthcare (defined as a method or technique used to enhance the adoption, implementation, and sustainability of a clinical program or practice), include either authorised healthcare practitioners or adults aged ≥ 18 years diagnosed with a musculoskeletal pain condition (21). Studies describing an implementation strategy or intervention but only evaluating the different treatments and not different implementation strategies or interventions of the same treatment were excluded. Studies that were non-experimental, involved animals, patients with serious pathology or patient populations below the age of 18 were excluded. No limitation on publication date and time periods were applied.

### Searches

The databases Medline via PubMed, EMBASE, Cochrane Central Register of Controlled Trials (CENTRAL), and Scopus were searched on the 22.02.2023 using the terms, “musculoskeletal Conditions”, “Implementation Strategies”, and “RCT”. Furthermore, relevant search terms were identified through manual searches, and standardised keywords and Medical Subject Headings (MeSH) were applied to ensure specificity for each individual database. An experienced librarian (JFJ) assisted in building the comprehensive literature search. Additionally, to ensure comprehensive coverage of the literature, the reference lists of eligible studies were reviewed to identify any potentially eligible studies using a forward citation search. See the full literature search in ***Additional file 2***.

### Study Selection

Covidence Software (Covidence Systematic Review Software, Veritas Health Innovation, Melbourne, Australia) was used in the selection process, and duplicates were removed from the initial search results. The selection process, which was performed by four independent authors (PBH, MSN, MEB and KDL), was divided into two phases. The first phase involved screening of titles and abstracts, and the second phase involved full-text screening. The screening was carried out by two independent reviewing authors for each study-in both phases (PBH, MSN, MEB or KDL). In case of conflicts between the independent authors or insufficient information from the title and abstract, the study was moved to full-text screening. In the second phase, the full text was examined by two authors to determine whether it met the eligibility criteria (PBH, MSN or MEB) (PBH, MSN or MEB). Conflicts in the full-text phase were resolved through a consensus discussion process involving three authors (PBH, MSN and MEB) and senior authors (MSR and KDL).

### Data Extraction

A purpose-built Excel sheet was used for data extraction. We collected and extracted data on study characteristics (study title, author, year, study design, country, clinical setting, sample size of HCPs and patients, and implementation strategy for intervention and control groups), and only findings of primary outcomes for HCPs, patients, and cost-effectiveness for each individual study. Three reviewers (PBH, MSN and MEB) performed data extraction from the included studies. For each study, there was a primary data extractor and one who checked for accuracy and completeness.

### Study Quality Assessment

To assess the risk of bias in individual studies, the Cochrane Risk of Bias tool 2 (A revised Cochrane risk of bias tool for randomised trials (RoB 2) was used (24). If the included studies were cluster randomised controlled trials, the RoB 2 tool for cluster-randomised trials was applied. Both tools assessed the risk of bias across five different domains, where RoB 2 for cluster-randomised trials divided the first domain into parts A and B. Each study was classified based on RoB 2 into three categories: high risk of bias, some concerns of risk of bias, or low risk of bias. Two independent reviewers (PBH, MSN or MEB) assessed the risk of bias, and conflicts were resolved through a consensus discussion process involving three authors (PBH, MSN and MEB) and senior authors (MSR and KDL).

### Data Synthesis and Presentation

Findings were presented descriptively. Two authors (PBH, MSN, MEB) independently categorised and mapped the implementation strategies in each study into implementation interventions and clusters using the nomenclature provided by Powell et al. 2015 and Waltz et al. 2015 (25,26). Discrepancies were resolved through a consensus discussion process involving three authors (PBH, MSN and MEB) while senior authors (MSR and KDL) were available if consensus could not be reached. The primary outcomes from each included study were condensed into a set of outcome domains through discussions. Three authors (PBH, MSN and MEB) mapped patient outcome domains using a core outcome set by Thompson et al. 2022 (27). Three authors (PBH, MSN and MEB) pragmatically mapped and formulated HCP outcome domains and outcomes of cost-effectiveness without using a core outcome set because none existed. However, inspiration for the HCP outcome domains was gathered from Proctor et al. 2011 (28). The process of creating the HCP outcome domains and the definitions of all outcome domains are presented in ***Additional file 3***. The findings of the primary outcomes in each study were used to create a tabular view of the effectiveness of different implementation interventions. All results concerning the primary outcomes in the included studies were divided into four categories of significance using probability values (p-values) (29,30). These four categories were defined as:

1. “Statistically significant positive effect in favour of intervention group” (S).
2. “No statistically significant positive effect in favour of intervention group” (NS).
3. “Mixed findings of statistically significant effect” (MS) for studies showing both S and NS classified within the same outcome domain.
4. “Unknown statistically significant effect” (US) for those who did not produce a p-value and showcased the results as descriptive statistics.

A statistically significant positive effect in favour of the intervention group was defined as a p-value of ≤0.05. However, due to some studies not presenting p-values, we interpreted odds ratios with 95% confidence intervals not including “1” and cost-effectiveness with 95% confidence intervals not including “0” as statistically significant.

## Results

### Study Selection

The systematic literature search resulted in 14,255 original results. Ten studies, not captured by the systematic literature search but found through manual searches, were also included (31–40). 14,196 studies were excluded during screening of titles and abstracts and 31 studies were excluded during screening of full texts. After screening 38 studies from 31 trials remained for final inclusion (31–68) (*See* ***Figure 1*** *for PRISMA Flow chart*).

**FIGURE 1:**
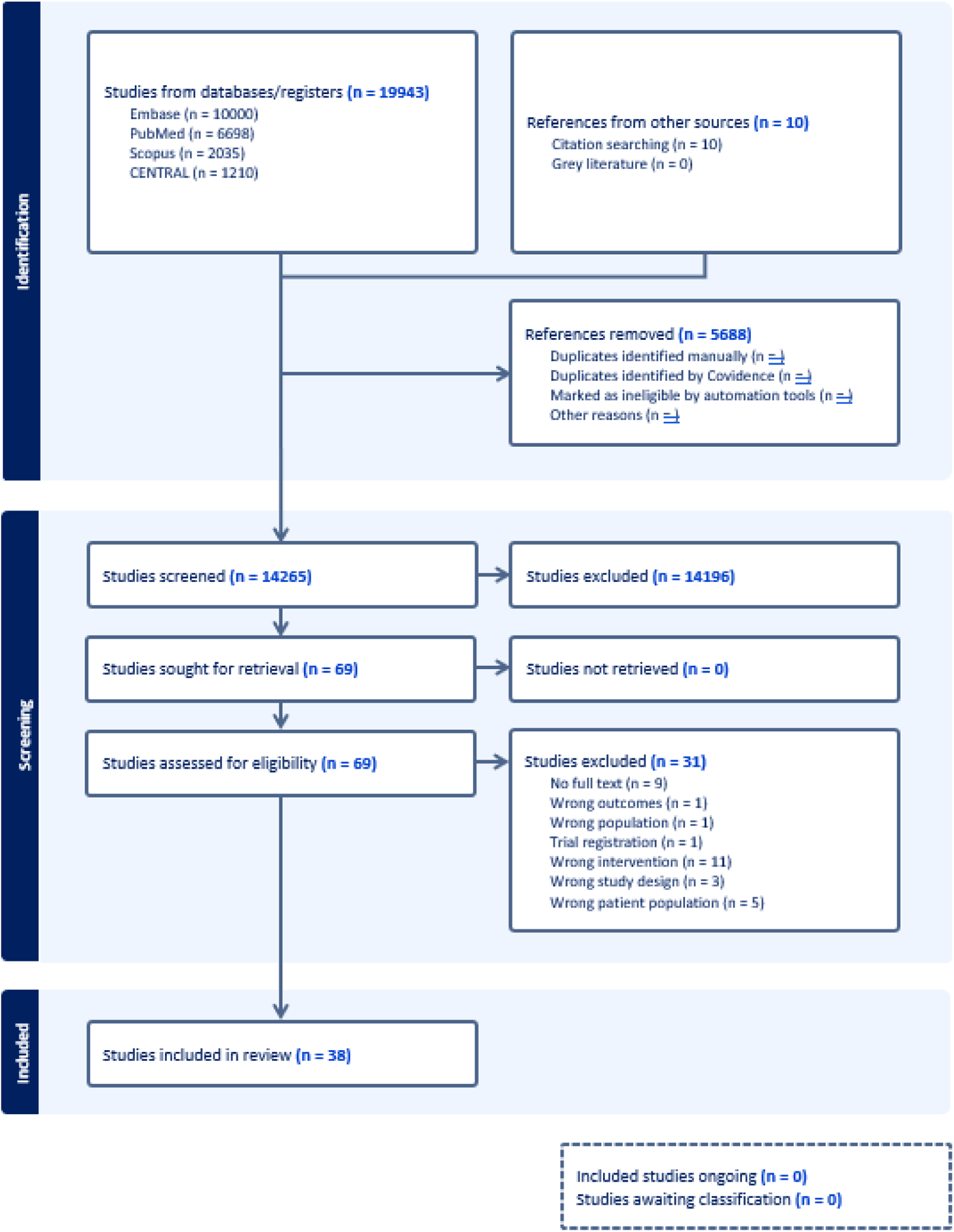
PRISMA FLOWCHART. The Preferred Reporting Items for Systematic Reviews and Meta-Analyses (PRISMA) Flowchart illustrating the screening and selection process including reasons for exclusion.

### Study Characteristics

The 38 included studies were published between 2001 and 2022. Of the 38 studies, six used a standard RCT design and 32 used a cluster RCT design, of which three utilised a stepped-wedge design. A full description of study characteristics including author, year, study design, sample size, clinical setting, country of origin, implementation strategies and interventions, primary outcomes and findings of the 38 included studies are summarised in ***Table 1***. ***Additional file 4*** presents ***Table 1*** in further detail including categorization of implementation interventions. Twenty-nine studies focused on measuring outcomes related to HCPs, ten studies on outcomes related to patients, and four studies on outcomes related to cost-effectiveness. A total of 13,203 HCPs were recruited in the 38 included studies. Twenty-one studies recruited general practitioners or physicians including family physicians, emergency physicians or primary care physicians (n=9558), 19 studies recruited physiotherapists (n=2194), one study involved osteopaths (n=598), three studies involved chiropractors (n=584), three studies involved nurses, rheumatologists, and spinal surgeons, respectively, but failed to report on the number of participants. Overall, five studies failed to report the number of recruited HCPs. A total of 30,320 patients were recruited in the 38 included studies. Thirty studies involved patients with LBP (n=26,774), four studies involved patients with neck pain (n=1460), five and four studies involved patients with knee pain and patients with hip pain, respectively, but failed to report the number of participants. Overall, 11 studies did not include patients or failed to report the number of recruited patients. Twelve of the included studies used data from the same RCT as other studies (35,41–44,52,54–56,59,65,66).

**TABLE 1:**
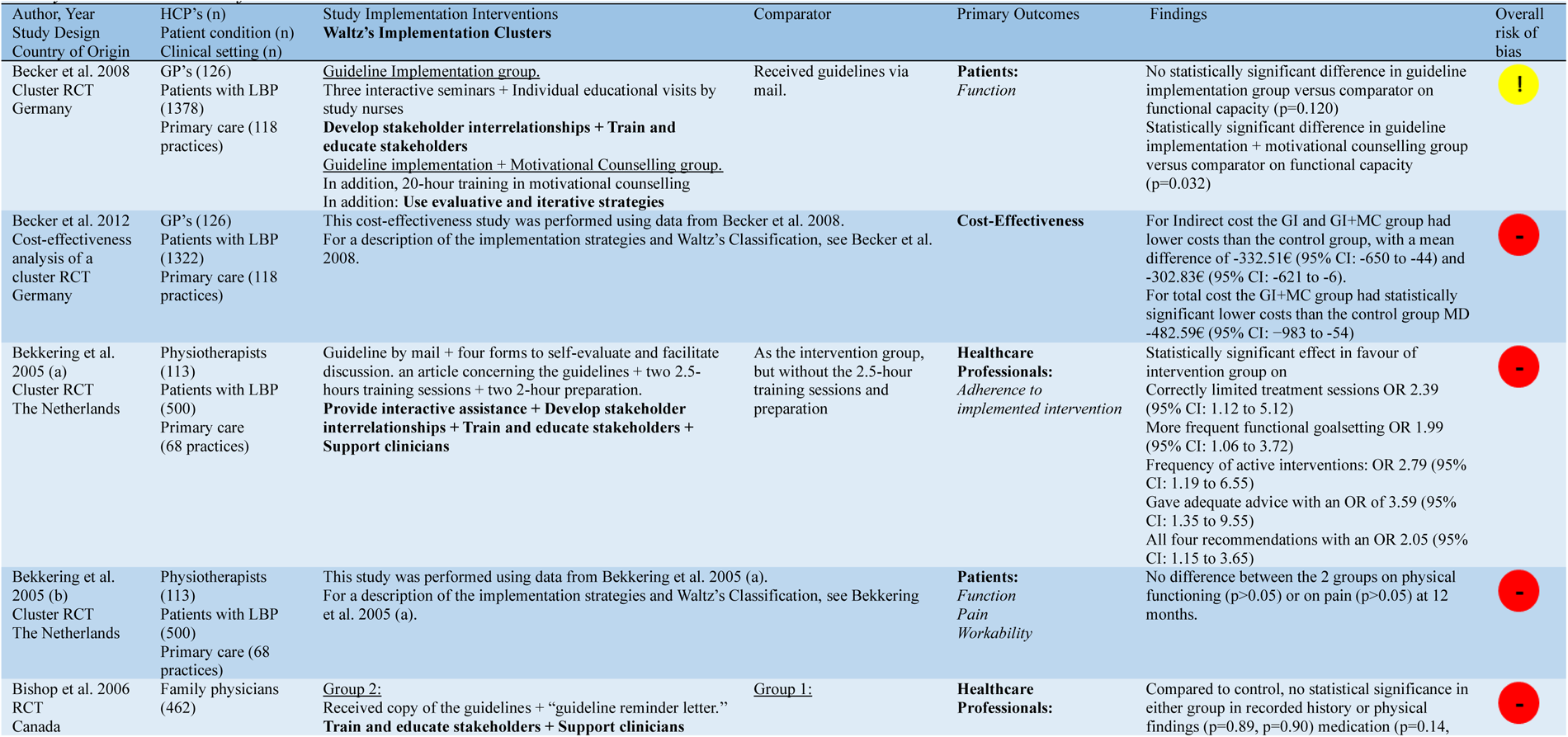
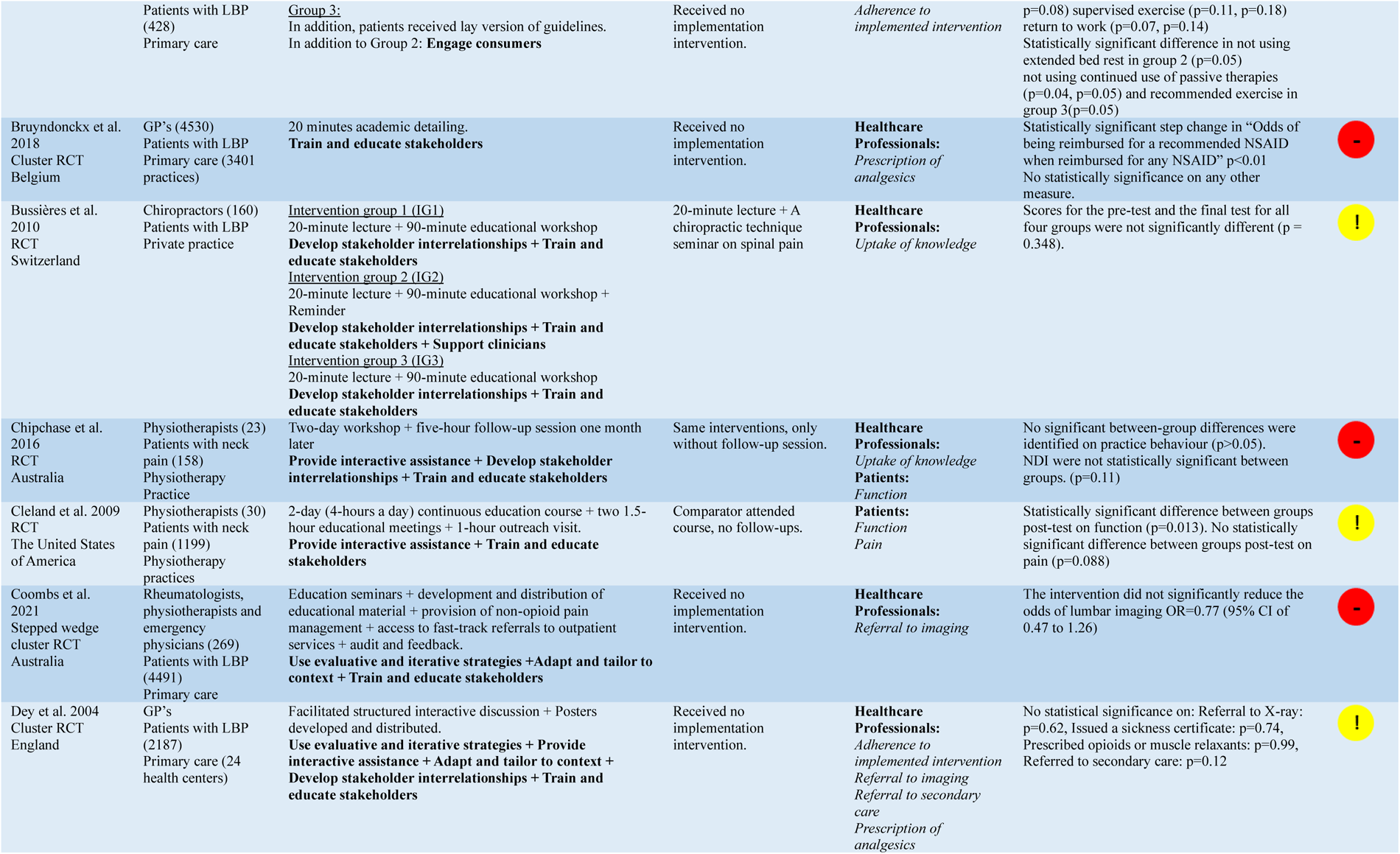
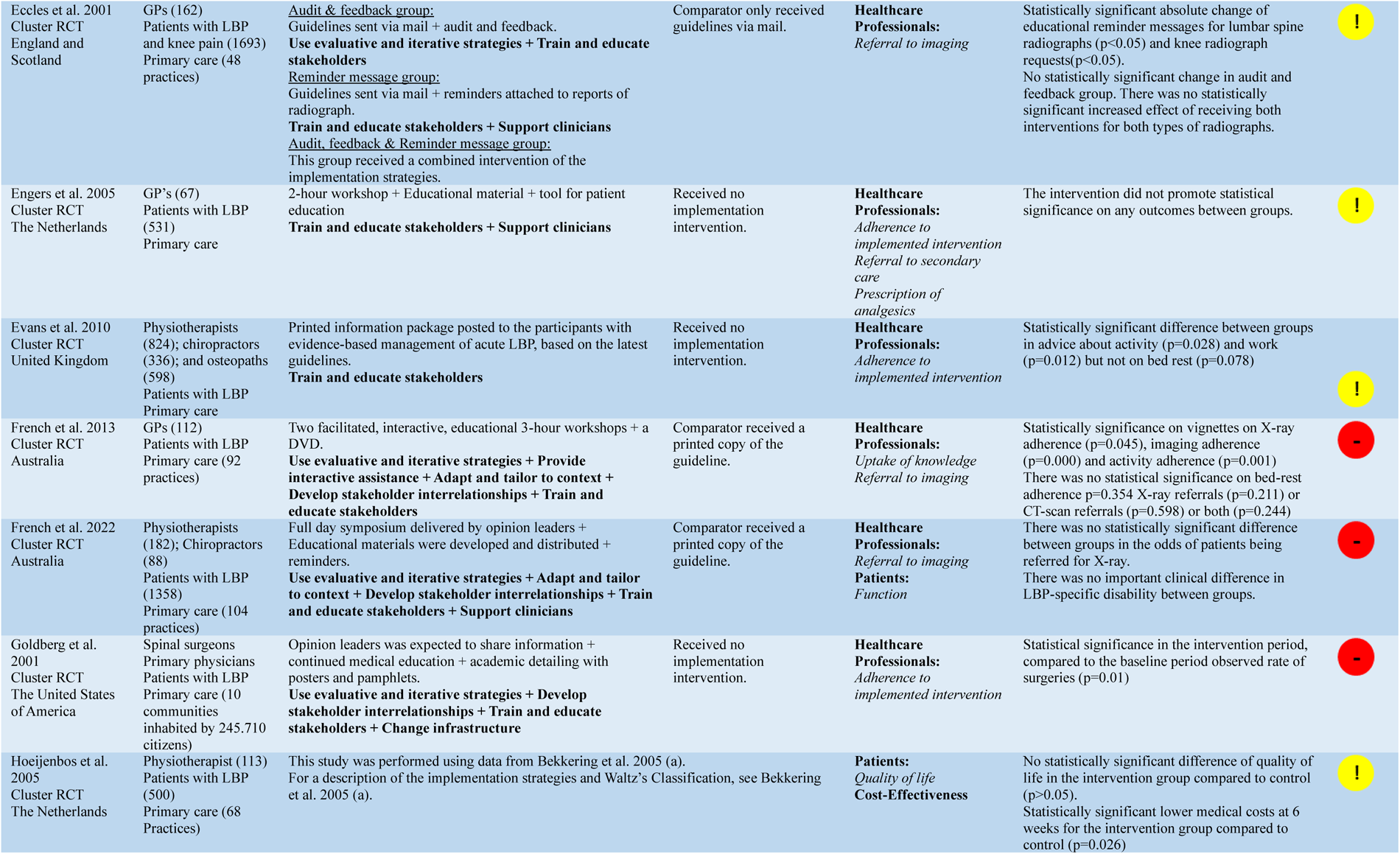
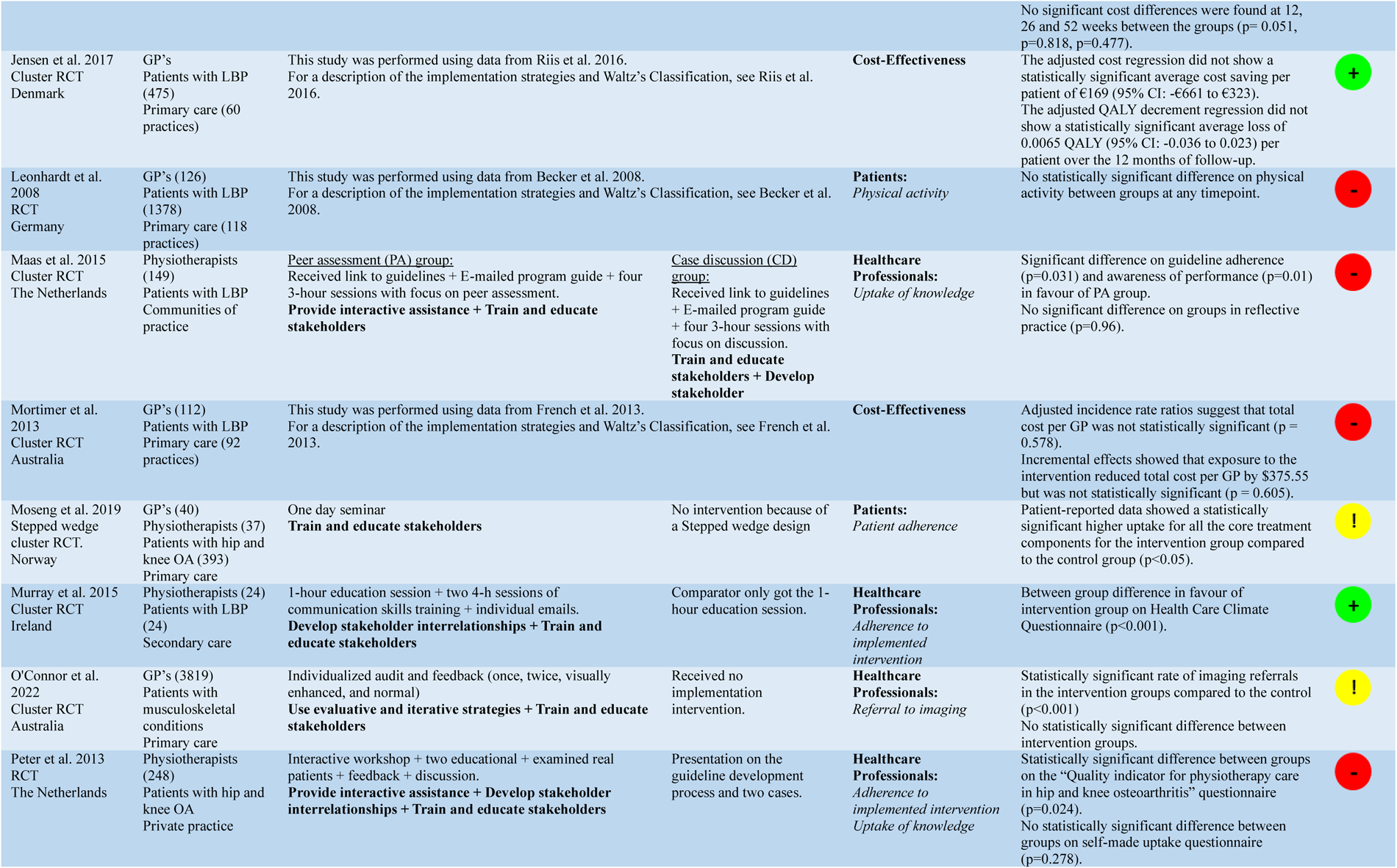
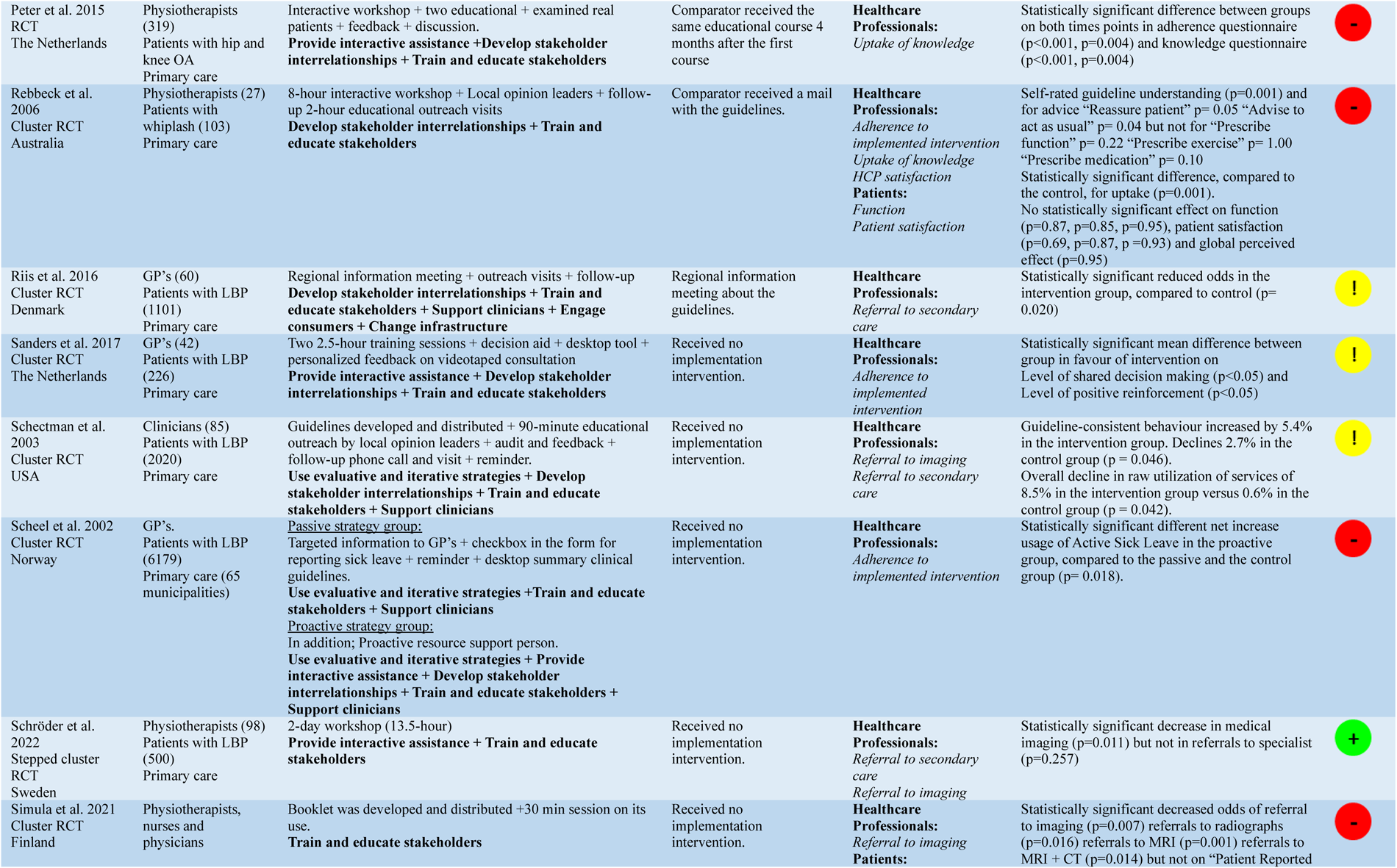
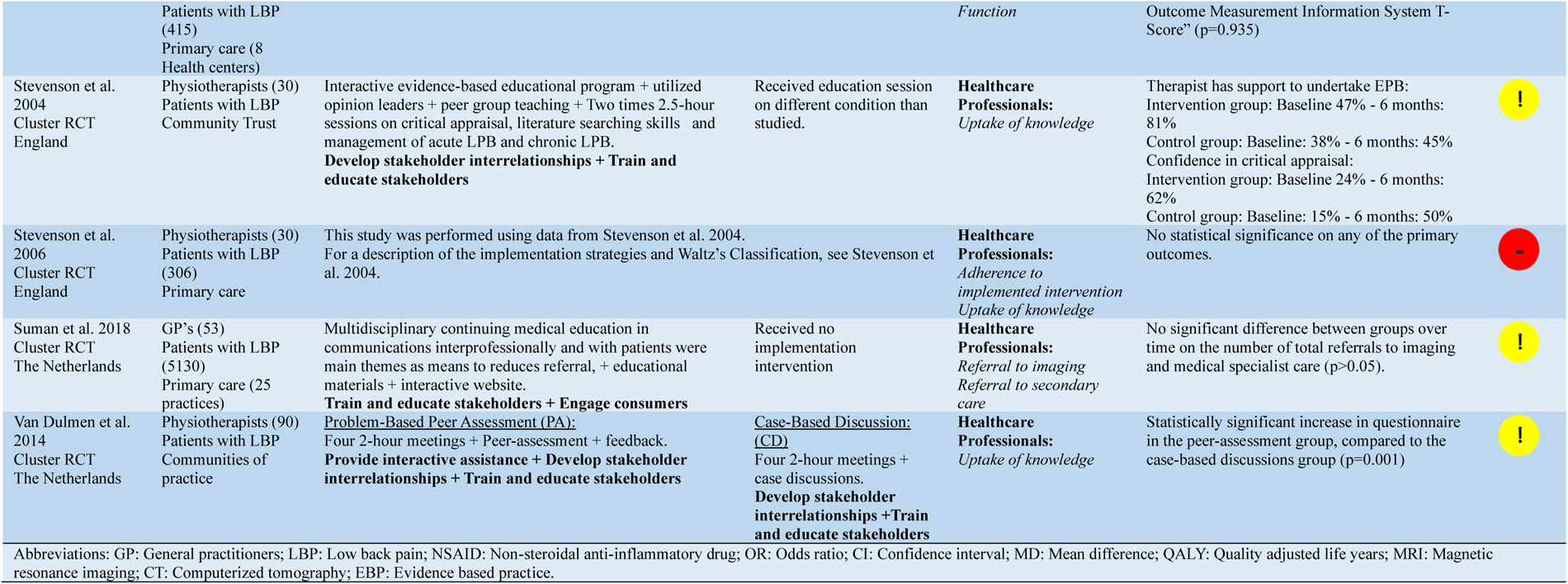
Study Characteristics of all Included Studies.

### Effect of Implementation Interventions

The categorization and mapping of the included studies, by implementation clusters and interventions combined with the outcome domains of primary outcomes for each implementation intervention, are presented in ***Figure 2***. ***Additional file 5*** presents the synthesis of the categorised included studies into implementation interventions and outcome domains with the corresponding references.

The most common intervention utilised in the literature is “Distribute educational materials” and 30 of the included studies utilised this implementation intervention. Among them, six out of ten studies assessing the outcome domain “Adherence to implemented intervention” found a statistically significant positive effect in favour of the intervention group. For the outcome domains “Referral to imaging”, “Referral to secondary care” and “Function” four out of ten studies, two out of six studies and one out of six studies found a statistically significant positive effect in favour of the intervention group, respectively.

Twenty-nine of the included studies utilised the implementation intervention “Conduct educational meetings”. Among them, seven out of nine studies assessing the outcome domain “Adherence to implemented intervention” found a statistically significant positive effect in favour of the intervention group. For the outcome domains “Uptake of knowledge”, and “Function”, three out of ten studies and two out of seven studies found a statistically significant positive effect in favour of the intervention group, respectively.

Twenty-two of the included studies utilised the implementation intervention “Conduct local consensus discussion”. Among them, five out of seven studies assessing the outcome domain “Adherence to implemented intervention” found a statistically significant positive effect in favour of the intervention group. For the outcome domain “Uptake of knowledge” two out of eight studies found a statistically significant positive effect in favour of the intervention group.

Twenty-one of the included studies utilised, the implementation intervention “Make training dynamic”. Among them, five out of seven studies assessing the outcome domain “Adherence to implemented intervention” found a statistically significant positive effect in favour of the intervention group. For the outcome domains “Uptake of knowledge”, and “Function”, three out of ten studies and one out of five studies found a statistically significant positive effect in favour of the intervention group, respectively.

Eighteen of the included studies utilised the implementation intervention “Conduct ongoing training”. Among these, two out of eight studies measuring the outcome domain “Uptake of knowledge” and two out of five studies measuring the outcome domain “Function” found a statistically significant positive effect in favour of the intervention group.

Sixteen of the included studies utilised the implementation intervention “Conduct educational outreach visits”. Two out of six studies measuring the outcome domain “Function” found a statistically significant positive effect in favour of the intervention group.

Four studies examined the cost-effectiveness. These studies produced inconsistent findings and employed varying outcome measures of effectiveness.

Lastly, across all outcome domains, several different outcome measures were utilised in the included studies See ***Additional file 4*** for the specific outcome measures utilised.

**FIGURE 2:**
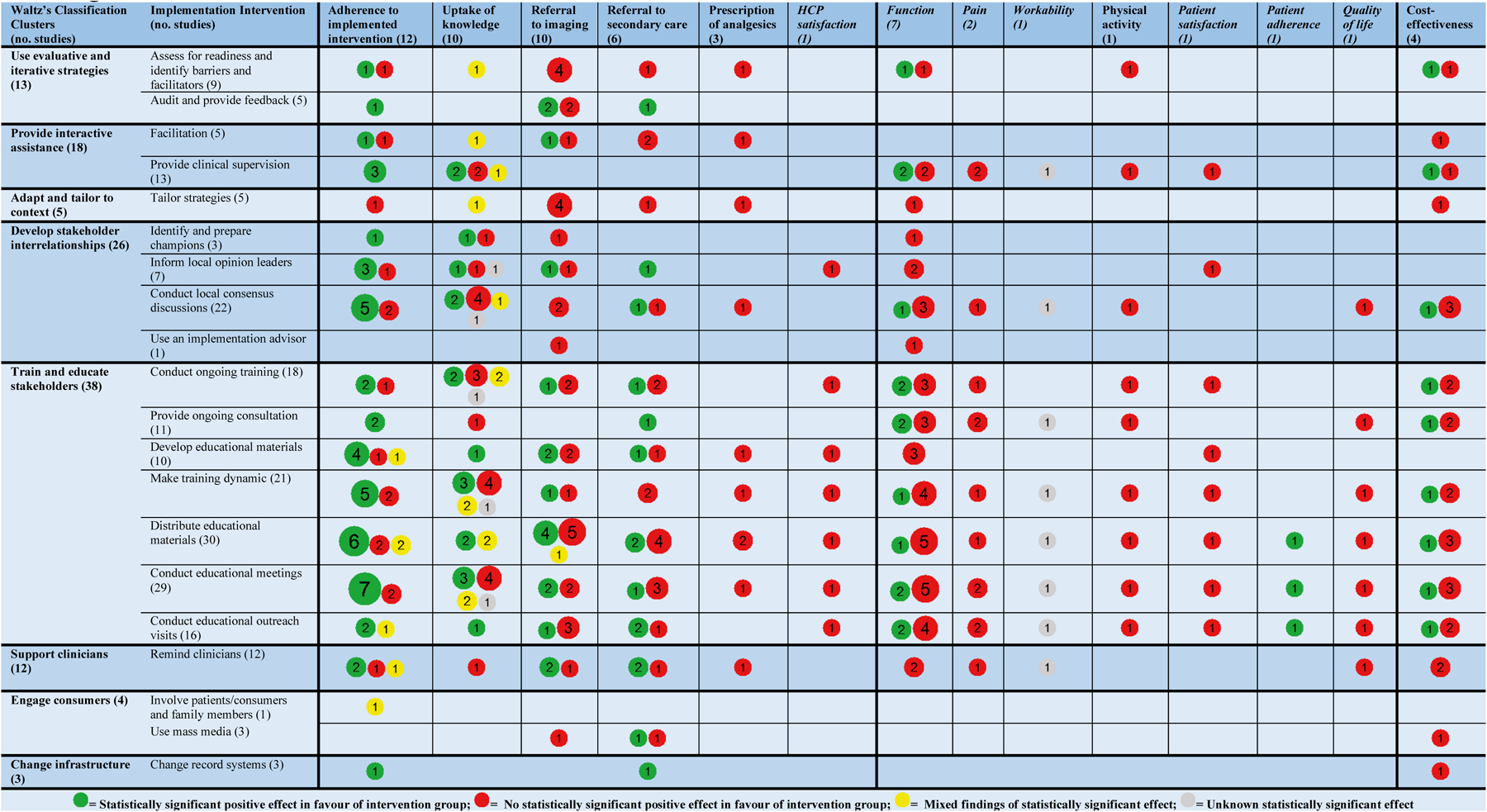
Mapping Implementation Interventions for Musculoskeletal Healthcare.

Map of significance illuminating the effect of implementation interventions for musculoskeletal healthcare. Types of implementation interventions are listed within the rows, and outcome domains are listed in the columns. The size of the circles and the number in the circles indicates the number of RCTs identified. A lack of RCTs results in missing circles in the corresponding fields.

### Study Quality Assessment

All 38 studies were assessed for risk of bias using RoB 2. Three studies were judged to have a low risk of bias, 16 studies to have some concerns about risk of bias, and 19 studies to have a high risk of bias. The overall results of the risk of bias assessment using RoB 2 is presented in ***Table 1***. ***Additional file 6*** illustrates a detailed version of the results of the risk of bias assessment for each RoB 2 domain.

## Discussion

### Summary of findings

This systematic review investigated the effectiveness of different implementation interventions in musculoskeletal healthcare and included 38 studies. Our synthesis of the findings from the included studies indicates that implementation interventions have diverse effects on HCP outcomes. Regarding studies measuring patient outcomes, implementation interventions may not lead to significant improvements. Conflicting findings were observed concerning cost-effectiveness outcomes, and we were unable to draw clear conclusions in this regard. Studies measuring the outcome domain “Adherence to the implemented intervention” may indicate a statistically significant positive effect in favour of the intervention group across most implementation interventions.

The most common implementation interventions utilised in the literature were “Distribute educational materials”, “Conduct educational meetings”, “Conduct local consensus discussion” and “Make training dynamic”. The majority of the studies utilising these interventions and measuring the outcome “Adherence to implemented intervention” found a statistically significant positive effect in favour of the intervention group.

Assessment of risk of bias using the RoB 2 tool revealed that most studies had either some concerns or a high risk of bias.

### Explanation of findings

The conflicting findings on HCP outcomes might be explained by significant methodological concerns such as the possibility of selection bias. It is likely that the HCPs who agreed to participate in the studies had extensive experience, continued education and were more inclined to be adherent with high quality of treatment and high baseline measurements, leaving less potential for improvement (31,32,38,39,48,49,51,52,59,61,63,66,67). Furthermore, the participating HCPs may have been more motivated with a positive attitude towards the implemented intervention implying a readiness to change (40,43,48–50,58,59). In addition to this, it is also possible that simply participating in a study and being observed may have resulted in greater adherence to the implemented intervention (48,58,59). These factors may have contributed to an increased effect of the implemented intervention in both the intervention and control groups across studies, thus obscuring the possible effect. The conflicting findings on patient outcomes and cost-effectiveness might equally be explained by the potential influence of selection bias, where HCPs in the control group possessed significant experience and interest in management of musculoskeletal conditions. This could potentially result in a higher likelihood of adhering to the implemented intervention with a high quality of treatment, leaving less potential for demonstrating a statistically significant difference between patient groups (32,35,40,44). Additionally, the absence of a significant effect may be attributed to unaccounted mediating confounders among patients, such as fear-avoidance and anxiety, as well as issues related to inclusion and substantial loss to follow-up (33,41,64). It is reasonable to suggest that the lack of effect on patient outcomes might, in part, contribute to the absence of cost-effectiveness (29,69). Only 20 of the 38 included studies provided a rationale for selecting the specific implementation strategy and interventions they examined (31–33,35–37,43,44,46,47,49–53,56,58,60,65,66). Despite providing a rationale for the choice of an implementation strategy and interventions, in many cases the rationale remained vague and not thoroughly reported. The absence of or vague rationales could potentially explain the conflicting findings observed in this review. Some literature illustrates the importance of providing a clear and well-thought rationale including exploring barriers and facilitators for changing practice behaviour, as this can affect the success of the implementation (26,70–73). In addition, the existing literature emphasises the importance of utilising implementation interventions targeting relevant mechanisms of change to address identified barriers and facilitators for behaviour change when selecting and tailoring of implementation strategies and interventions(74,75). Failing to tailor the implementation strategies and interventions towards relevant barriers and facilitators for behaviour change could result in random findings with small to moderate effects as observed in this review. Due to this it is recommendable that future research thoroughly describes the implementation strategy and provide an underlying rationale in detail.

### Agreements and Disagreements with Existing Literature

All single implementation interventions in this review showed various effects across all outcome domains, yet there seems to be a tendency towards a statistically significant effect in favour of the intervention group on the HCP outcome domain “Adherence to implemented intervention” for several implementation interventions. These findings align with existing literature within musculoskeletal healthcare indicating no single implementation intervention consistently outperform others across outcome domains (29,76–80). Similar findings are seen when looking at implementation interventions in a wider perspective within healthcare with implementation interventions such as providing audits and feedback, using local opinion leaders, using educational materials and using educational meetings and workshops potentially contributing to positive outcomes related to HCPs but not patients (81–84). This means that it might be recommendable to consider the following implementation interventions “Conduct local consensus discussions”, “Make training dynamic”, “Distribute educational materials“, “Conduct educational meetings”, “Audit and feedback” and “Using local opinion leaders” when creating an implementation strategy aiming at HCP behaviour change (81–84). Our results indicate a lack of positive outcomes in favour of the intervention group for patient outcome domains, which is consistent with findings from other reviews in musculoskeletal healthcare and broader healthcare implementation studies (29,67,76–78,80–85). This suggests that we currently lack effective implementation interventions to yield better patient outcomes. In addition, it could be speculated that the quality and efficacy of some of the implemented interventions and utilised measurements might be inadequate for changing and measuring patient outcomes. Our results concerning cost-effectiveness from four studies showed conflicting findings across implementation interventions. However, findings from existing literature show similar conflicting results, which could be explained by heterogeneous studies with different types of implementation interventions and outcomes (69,81). Due to this we, as well as other research, suggest that adding a cost-effectiveness analysis to a study concerning implementation interventions to demonstrate feasibility and contribute to more evidence regarding cost-effectiveness (67).

### Strength and Limitations

To our knowledge, this systematic review is the largest focusing on RCTs concerning implementation interventions in musculoskeletal healthcare, which is a considerable strength of this systematic review. Furthermore, the wide search strategy, which was developed in collaboration with an experienced librarian, yielded a much more comprehensive result than previous reviews on the subject (29,67,76–78,80). Despite this, we identified further 10 studies by manually searching, which showcase the substantial heterogeneity in both methodology and nomenclature in the field of implementation science. Our study has several limitations; we utilised the definition and nomenclature of implementation interventions as described in Powell et al. 2015 (25). However, a substantial amount of the definitions remain vague, and as such the classification is based on an individual assessment. This is equally in part to the poor description of implementation strategies and interventions in most of the included studies, and as such we were forced to make assumptions and individual interpretations in many cases. These challenges will undeniably result in discrepancies between reviews, as for instance in Goorts et al. 2021 there is a discordance in identifying and classifying interventions compared to ours (29). Therefore, future research should utilise standardised methodology and nomenclature to ensure consensus.

To our knowledge, this is the first review seeking to clarify which implementation interventions are effective within musculoskeletal healthcare. To do that, we decided to dissect the included studies’ implementation strategies into implementation interventions and classify these. However, in most studies, more than one implementation intervention could be identified, and it could be speculated that the effect sizes are due to multiple specific implementation interventions being used in conjunction with each other, which is a factor this review has not accounted for. Substantial heterogeneity was observed in the included studies. For example, when measuring adherence to the implemented intervention or patient functioning, there was hardly ever the same outcome measure appearing twice. To overcome this heterogeneity all primary outcomes were condensed into three sets of outcome domains. For patient outcomes we attempted to utilise a core outcome set for exercise and physical activity interventions for musculoskeletal disorders (27). However, not all patient outcomes were a core outcome according to Thompson et al. 2022, despite this, we chose to include these as domains regardless. This core outcome set was not in complete agreement with this study as we were not investigating treatments specifically consisting of exercise and physical activity, but to our knowledge nothing better existed. A limitation of this study was that to our knowledge no core outcome sets exist for HCP outcomes and cost-effectiveness. Therefore, to invent our own outcome domains will not be as generalizable to other studies as utilising an existing core outcome set. However, this domainisation of outcomes had the advantages that it made it possible to homogenise and synthesise the various heterogeneous outcomes of the included studies. Furthermore, we highlighted some implementation interventions to be more effective than others. These findings were based solely on observations of our synthesis, and as such it is important to note that these findings are not based on sample size, power or statistical effect sizes. Equally, no risk of bias comparisons of the studies could suggest a pattern. Despite this, our findings are in line with pre-existing literature (81–84).

## Conclusion

This systematic review offers an extensive overview of which individual implementation interventions that may be effective in musculoskeletal healthcare. Our data suggests that the implementation interventions “Conduct local consensus discussions”, “Make training dynamic”, “Distribute educational materials” and “Conduct educational meetings” may have a tendency towards a statistically significant positive effect in favour of the intervention group amongst HCPs concerning the outcome domain “Adherence to implemented intervention”. For the remaining HCP outcome domains and the cost-effectiveness outcome domain the effects are unknown across all implementation interventions due to discrepancies between studies. For patient outcome domains, it appears that no implementation interventions yields a statistically significant positive effect in favour of the intervention group. Most of the included studies were determined to have either some concerns or high risk of bias and had a high methodological heterogeneity. These results should therefore be interpreted with caution. For the field of implementation science to draw stronger conclusions and advance our knowledge about implementation strategies and interventions, it is important to conduct high-quality studies with detailed reporting of the methodology, including a comprehensive description of implementation strategies and interventions as well, as a well-defined rationale while utilising a standardised nomenclature. Additionally, it is important to allow for the investigation of behaviour change mechanisms to draw stronger conclusions and contribute to the advancement of knowledge in the field.

## Supporting information

PRISMA checklist

Literature search

Definition of outcome domains

Full version of table 1

Synthesis of results

Risk of Bias

## Data Availability

All data produced in the present study are available upon reasonable request to the authors.

## Declarations

### Ethics approval and consent to participate

Not applicable.

### Consent for publication

Not applicable.

### Availability of data and materials

All data generated during this study are included either within the text or as an additional file.

## Authors’ Contributions

All authors drafted the first protocol. PBH, MB, MSN, JFJ and KDL drafted the literature searches. PBH, MB, MSN, and KDL conducted the screening of all studies. PBH, MB, and MSN extracted all data. PBH, MB, MSN, MSR, and KDL participated in the data analysis. PBH, MB, and MSN, drafted the first draft and first review was made by MSR and KDL. Finally, all authors have thoroughly reviewed and approved the final submitted version of the manuscript.

## Acknowledgements

Not applicable.

## Contributions to the literature

⍰ This systematic review provides an overview of implementation interventions and the statistically significant positive effect in favour of the intervention groups within musculoskeletal healthcare.
⍰ The effect of implementation interventions on most outcome domains varies substantially, however, there seem to be a potential positive statistically significant effect of some implementation interventions for outcomes measuring the HCP outcome domain “Adherence to implemented intervention”.
⍰ This systematic review highlights the poor quality of trials found in the literature and advise future authors to extensively describe the implementation strategies and interventions and the rationale hereof in detail and utilising a standardised nomenclature to ensure consensus.

## References

1. Cieza A, Causey K, Kamenov K, Hanson SW, Chatterji S, Vos T. Global estimates of the need for rehabilitation based on the Global Burden of Disease study 2019: a systematic analysis for the Global Burden of Disease Study 2019. The Lancet. 2020 Dec;396(10267):2006–17.

2. Vos T, Lim SS, Abbafati C, Abbas KM, Abbasi M, Abbasifard M, et al. Global burden of 369 diseases and injuries in 204 countries and territories, 1990–2019: a systematic analysis for the Global Burden of Disease Study 2019. The Lancet. 2020 Oct;396(10258):1204–22.

3. Safiri S, Kolahi A, Cross M, Hill C, Smith E, Carson[Chahhoud K, et al. Prevalence, Deaths, and Disability[Adjusted Life Years Due to Musculoskeletal Disorders for 195 Countries and Territories 1990–2017. Arthritis & Rheumatology. 2021 Apr;73(4):702–14.

4. Gaskin DJ, Richard P. The economic costs of pain in the United States. J Pain. 2012 Aug;13(8):715–24.

5. Mairey I, Rosenkilde S, Klitgaard MB, Thygesen LC. Sygdomsbyrden i Danmark: sygdomme [Internet]. Version 2.0. Statens Institut for Folkesundhed, Syddansk Universitet. Sygdomsbyrden i Danmark – sygdomme. København: Sundhedsstyrelsen; 2022; 2023 [cited 2023 May 25]. Available from: https://www.sst.dk/-/media/Udgivelser/2023/Sygdomsbyrden-2023/Sygdomme-Sygdomsbyrden-2023.ashx

6. Bevan S. Economic impact of musculoskeletal disorders (MSDs) on work in Europe. Best Practice & Research Clinical Rheumatology. 2015 Jun;29(3):356–73.

7. Brownlee S, Chalkidou K, Doust J, Elshaug AG, Glasziou P, Heath I, et al. Evidence for overuse of medical services around the world. Lancet. 2017 Jul 8;390(10090):156–68.

8. Chou R, Fu R, Carrino JA, Deyo RA. Imaging strategies for low-back pain: systematic review and meta-analysis. Lancet. 2009 Feb 7;373(9662):463–72.

9. Jenkins HJ, Downie AS, Maher CG, Moloney NA, Magnussen JS, Hancock MJ. Imaging for low back pain: is clinical use consistent with guidelines? A systematic review and meta-analysis. Spine J. 2018 Dec;18(12):2266–77.

10. Maher CG, O’Keeffe M, Buchbinder R, Harris IA. Musculoskeletal healthcare: Have we over-egged the pudding? Int J Rheum Dis. 2019 Nov;22(11):1957–60.

11. Morgan DJ, Dhruva SS, Coon ER, Wright SM, Korenstein D. 2019 Update on Medical Overuse: A Review. JAMA Intern Med. 2019 Nov 1;179(11):1568–74.

12. Müskens JLJM, Kool RB, van Dulmen SA, Westert GP. Overuse of diagnostic testing in healthcare: a systematic review. BMJ Qual Saf. 2022 Jan;31(1):54–63.

13. Zadro J, O’Keeffe M, Maher C. Do physical therapists follow evidence-based guidelines when managing musculoskeletal conditions? Systematic review. BMJ Open. 2019 Oct;9(10):e032329.

14. Malhotra A, Maughan D, Ansell J, Lehman R, Henderson A, Gray M, et al. Choosing Wisely in the UK: the Academy of Medical Royal Colleges’ initiative to reduce the harms of too much medicine. BMJ. 2015 May 12;350(may12 7):h2308–h2308.

15. Levinson W, Born K, Wolfson D. Choosing Wisely Campaigns: A Work in Progress. JAMA. 2018 May 15;319(19):1975.

16. Malling B, Høffer M, Raft CF, Axelsen S. The Danish Choosing Wisely concept. Dan Med J. 2021 Sep 28;68(10):A11200889.

17. Cassel CK. Choosing Wisely: Helping Physicians and Patients Make Smart Decisions About Their Care. JAMA. 2012 May 2;307(17):1801.

18. Grol R, Grimshaw J. From best evidence to best practice: effective implementation of change in patients’ care. Lancet. 2003 Oct 11;362(9391):1225–30.

19. Morris ZS, Wooding S, Grant J. The answer is 17 years, what is the question: understanding time lags in translational research. J R Soc Med. 2011 Dec;104(12):510–20.

20. Bauer MS, Kirchner J. Implementation science: What is it and why should I care? Psychiatry Research. 2020 Jan;283:112376.

21. Curran GM, Bauer M, Mittman B, Pyne JM, Stetler C. Effectiveness-implementation Hybrid Designs: Combining Elements of Clinical Effectiveness and Implementation Research to Enhance Public Health Impact. Medical Care. 2012 Mar;50(3):217–26.

22. Powell BJ, Beidas RS, Lewis CC, Aarons GA, McMillen JC, Proctor EK, et al. Methods to Improve the Selection and Tailoring of Implementation Strategies. J Behav Health Serv Res. 2017 Apr;44(2):177–94.

23. Rowe R, McDaid D. Implementation: the need for a contextual approach to the implementation of musculoskeletal guidelines. Best Practice & Research Clinical Rheumatology. 2007 Feb;21(1):205–19.

24. Sterne JAC, Savović J, Page MJ, Elbers RG, Blencowe NS, Boutron I, et al. RoB 2: a revised tool for assessing risk of bias in randomised trials. BMJ. 2019 Aug 28;l4898.

25. Powell BJ, Waltz TJ, Chinman MJ, Damschroder LJ, Smith JL, Matthieu MM, et al. A refined compilation of implementation strategies: results from the Expert Recommendations for Implementing Change (ERIC) project. Implementation Sci. 2015 Dec;10(1):21.

26. Waltz TJ, Powell BJ, Matthieu MM, Damschroder LJ, Chinman MJ, Smith JL, et al. Use of concept mapping to characterize relationships among implementation strategies and assess their feasibility and importance: results from the Expert Recommendations for Implementing Change (ERIC) study. Implementation Sci. 2015 Dec;10(1):109.

27. Thompson A, Ross M, Tanzila P, Deb H, Matthew K, Sionnadh M. A Core Outcome Set for Exercise and Physical Activity Interventions for Musculoskeletal Disorders. J Musculoskelet Disord Treat [Internet]. 2022 Dec 31 [cited 2023 Mar 28];8(4). Available from: https://clinmedjournals.org/articles/jmdt/journal-of-musculoskeletal-disorders-and-treatment-jmdt-8-119.php?jid=jmdt

28. Proctor E, Silmere H, Raghavan R, Hovmand P, Aarons G, Bunger A, et al. Outcomes for Implementation Research: Conceptual Distinctions, Measurement Challenges, and Research Agenda. Adm Policy Ment Health. 2011 Mar;38(2):65–76.

29. Goorts K, Dizon J, Milanese S. The effectiveness of implementation strategies for promoting evidence informed interventions in allied healthcare: a systematic review. BMC Health Serv Res. 2021 Dec;21(1):241.

30. Morrissey D, Cotchett M, Said J’Bari A, Prior T, Griffiths IB, Rathleff MS, et al. Management of plantar heel pain: a best practice guide informed by a systematic review, expert clinical reasoning and patient values. Br J Sports Med. 2021 Oct;55(19):1106–18.

31. Bussières AE, Laurencelle L, Peterson C. Diagnostic Imaging Guidelines Implementation Study for Spinal Disorders: A Randomized Trial with Postal Follow-ups*. Journal of Chiropractic Education. 2010 Apr 1;24(1):2–18.

32. Chipchase LS, Cavaleri R, Jull G. Can a professional development workshop with follow-up alter practitioner behaviour and outcomes for neck pain patients? A randomised controlled trial. Man Ther. 2016 Sep;25:87–93.

33. Cleland JA, Fritz JM, Brennan GP, Magel J. Does Continuing Education Improve Physical Therapists’ Effectiveness in Treating Neck Pain? A Randomized Clinical Trial. 2009;

34. Eccles M, Steen N, Grimshaw J, Thomas L, McNamee P, Soutter J, et al. Effect of audit and feedback, and reminder messages on primary-care radiology referrals: a randomised trial. The Lancet. 2001 May;357(9266):1406–9.

35. Hoeijenbos M, Bekkering T, Lamers L, Hendriks E, van Tulder M, Koopmanschap M. Cost-effectiveness of an active implementation strategy for the Dutch physiotherapy guideline for low back pain. Health Policy. 2005 Dec;75(1):85–98.

36. Maas MJM, van der Wees PJ, Braam C, Koetsenruijter J, Heerkens YF, van der Vleuten CPM, et al. An innovative peer assessment approach to enhance guideline adherence in physical therapy: single-masked, cluster-randomized controlled trial. Phys Ther. 2015 Apr;95(4):600–12.

37. O’Connor DA, Glasziou P, Maher CG, McCaffery KJ, Schram D, Maguire B, et al. Effect of an Individualized Audit and Feedback Intervention on Rates of Musculoskeletal Diagnostic Imaging Requests by Australian General Practitioners: A Randomized Clinical Trial. JAMA. 2022 Sep 6;328(9):850.

38. Peter W, van der Wees PJ, Verhoef J, de Jong Z, van Bodegom-Vos L, Hilberdink WKHA, et al. Effectiveness of an interactive postgraduate educational intervention with patient participation on the adherence to a physiotherapy guideline for hip and knee osteoarthritis: a randomised controlled trial. Disabil Rehabil. 2015;37(3):274–82.

39. Peter WF, van der Wees PJ, Verhoef J, de Jong Z, van Bodegom-Vos L, Hilberdink WKHA, et al. Postgraduate education to increase adherence to a Dutch physiotherapy practice guideline for hip and knee OA: a randomized controlled trial. Rheumatology (Oxford). 2013 Feb;52(2):368–75.

40. Rebbeck T, Maher CG, Refshauge KM. Evaluating two implementation strategies for whiplash guidelines in physiotherapy: A cluster-randomised trial. Australian Journal of Physiotherapy. 2006;52(3):165–74.

41. Becker A, Leonhardt C, Kochen MM, Keller S, Wegscheider K, Baum E, et al. Effects of Two Guideline Implementation Strategies on Patient Outcomes in Primary Care: A Cluster Randomized Controlled Trial. Spine. 2008 Mar;33(5):473–80.

42. Becker A, Held H, Redaelli M, Chenot JF, Leonhardt C, Keller S, et al. Implementation of a Guideline for Low Back Pain Management in Primary Care: A Cost-Effectiveness Analysis. Spine. 2012 Apr;37(8):701–10.

43. Bekkering GE, Hendriks HJM, van Tulder MW, Knol DL, Hoeijenbos M, Oostendorp R a. B, et al. Effect on the process of care of an active strategy to implement clinical guidelines on physiotherapy for low back pain: a cluster randomised controlled trial. Qual Saf Health Care. 2005 Apr;14(2):107–12.

44. Bekkering GE, Van Tulder MW, Hendriks EJ, Koopmanschap MA, Knol DL, Bouter LM, et al. Implementation of Clinical Guidelines on Physical Therapy for Patients With Low Back Pain: Randomized Trial Comparing Patient Outcomes After a Standard and Active Implementation Strategy. Physical Therapy. 2005 Jun 1;85(6):544–55.

45. Bishop PB, Wing PC. Knowledge transfer in family physicians managing patients with acute low back pain: a prospective randomized control trial. The Spine Journal. 2006 May;6(3):282–8.

46. Bruyndonckx R, Verhoeven V, Anthierens S, Cornelis K, Ackaert K, Gielen B, et al. The implementation of academic detailing and its effectiveness on appropriate prescribing of pain relief medication: a real-world cluster randomized trial in Belgian general practices. Implementation Sci. 2018 Dec;13(1):6.

47. Coombs DM, Machado GC, Richards B, Needs C, Buchbinder R, Harris IA, et al. Effectiveness of a multifaceted intervention to improve emergency department care of low back pain: a stepped-wedge, cluster-randomised trial. BMJ Qual Saf. 2021 Oct;30(10):825–35.

48. Dey P, Simpson CWR, Collins SI, Hodgson G, Dowrick CF, Simison AJM, et al. Implementation of RCGP guidelines for acute low back pain: a cluster randomised controlled trial. Br J Gen Pract. 2004 Jan;54(498):33–7.

49. Engers AJ, Wensing M, van Tulder MW, Timmermans A, Oostendorp RAB, Koes BW, et al. Implementation of the Dutch Low Back Pain Guideline for General Practitioners: A Cluster Randomized Controlled Trial. Spine. 2005 Mar;30(6):559–600.

50. Evans DW, Breen AC, Pincus T, Sim J, Underwood M, Vogel S, et al. The Effectiveness of a Posted Information Package on the Beliefs and Behavior of Musculoskeletal Practitioners: The UK Chiropractors, Osteopaths, and Musculoskeletal Physiotherapists Low Back Pain ManagemENT (COMPLeMENT) Randomized Trial. Spine. 2010 Apr;35(8):858–66.

51. French SD, O’Connor DA, Green SE, Page MJ, Mortimer DS, Turner SL, et al. Improving adherence to acute low back pain guideline recommendations with chiropractors and physiotherapists: the ALIGN cluster randomised controlled trial. Trials. 2022 Dec;23(1):142.

52. French SD, McKenzie JE, O’Connor DA, Grimshaw JM, Mortimer D, Francis JJ, et al. Evaluation of a Theory-Informed Implementation Intervention for the Management of Acute Low Back Pain in General Medical Practice: The IMPLEMENT Cluster Randomised Trial. Gagnier JJ, editor. PLoS ONE. 2013 Jun 13;8(6):e65471.

53. Goldberg HI, Deyo RA, Taylor VM, Cheadle AD, Conrad DA, Loeser JD, et al. Can evidence change the rate of back surgery? A randomized trial of community-based education. Eff Clin Pract. 2001;4(3):95–104.

54. Jensen CE, Riis A, Petersen KD, Jensen MB, Pedersen KM. Economic evaluation of an implementation strategy for the management of low back pain in general practice. Pain. 2017 May;158(5):891–9.

55. Leonhardt C, Keller S, Chenot JF, Luckmann J, Basler HD, Wegscheider K, et al. TTM-based motivational counselling does not increase physical activity of low back pain patients in a primary care setting—A cluster-randomized controlled trial. Patient Education and Counseling. 2008 Jan;70(1):50–60.

56. Mortimer D, French SD, McKenzie JE, O′Connor DA, Green SE. Economic Evaluation of Active Implementation versus Guideline Dissemination for Evidence-Based Care of Acute Low-Back Pain in a General Practice Setting. Manchikanti L, editor. PLoS ONE. 2013 Oct 11;8(10):e75647.

57. Moseng T, Dagfinrud H, Østerås N. Implementing international osteoarthritis guidelines in primary care: uptake and fidelity among health professionals and patients. Osteoarthritis and Cartilage. 2019 Aug;27(8):1138–47.

58. Murray A, Hall AM, Williams GC, McDonough SM, Ntoumanis N, Taylor IM, et al. Effect of a Self-Determination Theory–Based Communication Skills Training Program on Physiotherapists’ Psychological Support for Their Patients With Chronic Low Back Pain: A Randomized Controlled Trial. Archives of Physical Medicine and Rehabilitation. 2015 May;96(5):809–16.

59. Riis A, Jensen CE, Bro F, Maindal HT, Petersen KD, Bendtsen MD, et al. A multifaceted implementation strategy versus passive implementation of low back pain guidelines in general practice: a cluster randomised controlled trial. Implementation Sci. 2016 Dec;11(1):143.

60. Sanders ARJ, Bensing JM, Essed MALU, Magnée T, de Wit NJ, Verhaak PFM. Does training general practitioners result in more shared decision making during consultations? Patient Education and Counseling. 2017 Mar;100(3):563–74.

61. Schectman JM, Schroth WS, Verme D, Voss JD. Randomized controlled trial of education and feedback for implementation of guidelines for acute low back pain. J Gen Intern Med. 2003 Oct;18(10):773–80.

62. Scheel IB, Hagen KB, Herrin J, Oxman AD. A Randomized Controlled Trial of Two Strategies to Implement Active Sick Leave for Patients With Low Back Pain.

63. Schröder K, Öberg B, Enthoven P, Hedevik H, Abbott A. Improved adherence to clinical guidelines for low back pain after implementation of the BetterBack model of care: A stepped cluster randomized controlled trial within a hybrid type 2 trial. Physiotherapy Theory and Practice. 2022 Mar 1;1–15.

64. Simula AS, Jenkins HJ, Hancock MJ, Malmivaara A, Booth N, Karppinen J. Patient education booklet to support evidence-based low back pain care in primary care – a cluster randomized controlled trial. BMC Fam Pract. 2021 Dec;22(1):178.

65. Stevenson K, Lewis M, Hay E. Do physiotherapists’ attitudes towards evidence-based practice change as a result of an evidence-based educational programme?: Can we change physiotherapist’s attitudes towards EBP? Journal of Evaluation in Clinical Practice. 2004 May;10(2):207–17.

66. Stevenson K, Lewis M, Hay E. Does physiotherapy management of low back pain change as a result of an evidence-based educational programme? J Eval Clin Pract. 2006 Jun;12(3):365–75.

67. Suman A, Schaafsma FG, van de Ven PM, Slottje P, Buchbinder R, van Tulder MW, et al. Effectiveness of a multifaceted implementation strategy compared to usual care on low back pain guideline adherence among general practitioners. BMC Health Serv Res. 2018 Dec;18(1):358.

68. van Dulmen SA, Maas M, Staal JB, Rutten G, Kiers H, Nijhuis-van der Sanden M, et al. Effectiveness of peer assessment for implementing a Dutch physical therapy low back pain guideline: cluster randomized controlled trial. Phys Ther. 2014 Oct;94(10):1396–409.

69. Jensen CE, Jensen MB, Riis A, Petersen KD. Systematic review of the cost-effectiveness of implementing guidelines on low back pain management in primary care: is transferability to other countries possible? BMJ Open. 2016 Jun;6(6):e011042.

70. Morris JH, Bernhardsson S, Bird ML, Connell L, Lynch E, Jarvis K, et al. Implementation in rehabilitation: a roadmap for practitioners and researchers. Disability and Rehabilitation. 2020 Oct 22;42(22):3265–74.

71. Nilsen P. Making sense of implementation theories, models and frameworks. Implementation Sci. 2015 Dec;10(1):53.

72. Nilsen P, Ingvarsson S, Hasson H, von Thiele Schwarz U, Augustsson H. Theories, models, and frameworks for de-implementation of low-value care: A scoping review of the literature. Implementation Research and Practice. 2020 Jan;1:263348952095376.

73. Weiner BJ. A theory of organizational readiness for change. Implementation Sci. 2009 Dec;4(1):67.

74. Lewis CC, Klasnja P, Powell BJ, Lyon AR, Tuzzio L, Jones S, et al. From Classification to Causality: Advancing Understanding of Mechanisms of Change in Implementation Science. Front Public Health. 2018 May 7;6:136.

75. Powell BJ, Fernandez ME, Williams NJ, Aarons GA, Beidas RS, Lewis CC, et al. Enhancing the Impact of Implementation Strategies in Healthcare: A Research Agenda. Front Public Health. 2019 Jan 22;7:3.

76. AL Zoubi FM, Menon A, Mayo NE, Bussières AE. The effectiveness of interventions designed to increase the uptake of clinical practice guidelines and best practices among musculoskeletal professionals: a systematic review. BMC Health Serv Res. 2018 Dec;18(1):435.

77. Fillipo R, Pruka K, Carvalho M, Horn ME, Moore J, Ramger B, et al. Does the implementation of clinical practice guidelines for low back and neck pain by physical therapists improve patient outcomes? A systematic review. Implement Sci Commun. 2022 Dec;3(1):57.

78. Mesner SA, Foster NE, French SD. Implementation interventions to improve the management of non-specific low back pain: a systematic review. BMC Musculoskelet Disord. 2016 Dec;17(1):258.

79. Suman A, Dikkers MF, Schaafsma FG, van Tulder MW, Anema JR. Effectiveness of multifaceted implementation strategies for the implementation of back and neck pain guidelines in health care: a systematic review. Implementation Sci. 2015 Dec;11(1):126.

80. van der Wees PJ, Jamtvedt G, Rebbeck T, de Bie RA, Dekker J, Hendriks EJM. Multifaceted strategies may increase implementation of physiotherapy clinical guidelines: a systematic review. Aust J Physiother. 2008;54(4):233–41.

81. Flodgren G, O’Brien MA, Parmelli E, Grimshaw JM. Local opinion leaders: effects on professional practice and healthcare outcomes. Cochrane Effective Practice and Organisation of Care Group, editor. Cochrane Database of Systematic Reviews [Internet]. 2019 Jun 24 [cited 2023 May 11];2019(6). Available from: http://doi.wiley.com/10.1002/14651858.CD000125.pub5

82. Forsetlund L, O’Brien MA, Forsén L, Mwai L, Reinar LM, Okwen MP, et al. Continuing education meetings and workshops: effects on professional practice and healthcare outcomes. Cochrane Effective Practice and Organisation of Care Group, editor. Cochrane Database of Systematic Reviews [Internet]. 2021 Sep 15 [cited 2023 May 11];2021(9). Available from: http://doi.wiley.com/10.1002/14651858.CD003030.pub3

83. 83. Giguère A, Zomahoun HTV, Carmichael PH, Uwizeye CB, Légaré F, Grimshaw JM, et al. Printed educational materials: effects on professional practice and healthcare outcomes. Cochrane Effective Practice and Organisation of Care Group, editor. Cochrane Database of Systematic Reviews [Internet]. 2020 Jul 31 [cited 2023 May 11];2020(8). Available from: http://doi.wiley.com/10.1002/14651858.CD004398.pub4

84. Ivers N, Jamtvedt G, Flottorp S, Young JM, Odgaard-Jensen J, French SD, et al. Audit and feedback: effects on professional practice and healthcare outcomes. Cochrane Effective Practice and Organisation of Care Group, editor. Cochrane Database of Systematic Reviews [Internet]. 2012 Jun 13 [cited 2023 May 10]; Available from: https://doi.wiley.com/10.1002/14651858.CD000259.pub3

85. O’Brien MA, Rogers S, Jamtvedt G, Oxman A, Odgaard-Jensen J, Kristoffersen DT, et al. Educational outreach visits: effects on professional practice and health care outcomes. CDSR [Internet]. 2007 Oct 17 [cited 2023 May 11]; Available from: http://doi.wiley.com/10.1002/14651858.CD000409.pub2

