## Supplementary material for "Effectiveness of Implementation Interventions in Musculoskeletal Healthcare: A Systematic Review": Literature search

### Additional file 2 - The Systematic Literature Search

| ***Table A1.1***  *Overview of the Results of the Literature Search from each Database* | | | |
| --- | --- | --- | --- |
| **Database** | **Interface** | **Results** | **Date** |
| PubMed | PubMed.gov | 6,698 | 22-02-2023 |
| Embase | Embase.com | 10,000 | 22-02-2023 |
| Scopus | Scopus.com | 2,035 | 22-02-2023 |
| Cochraine (CENTRAL) | cochranelibrary.com/central | 1,210 | 22-02-2023 |
| All  After removal of duplicates |  | 19,943  14,255 |  |

#### Medline via PubMed

| ***Table A1.2***  *The literature search from Medline via PubMed* | | | | |
| --- | --- | --- | --- | --- |
| **Search number** | **Search subjects** | **Search** | **Hits** | **Date** |
| **#1** | Musculoskeletal Conditions | (“Musculoskeletal*” OR "Musculoskeletal System"[MeSH] OR "Musculoskeletal Diseases"[Mesh] OR "Musculoskeletal and Neural Physiological Phenomena"[MeSH] OR "Myofascial pain syndromes"[MeSH] OR "Myofascial pain" OR "Musculoskeletal Pain"[MeSH] OR “Arthralgia” OR “Arthralgia”[MeSH] OR “Bursit*” OR “Tendin*” OR “Tendon*” OR “Myalgia” OR “Joint pain” OR “Muscle pain” OR “Soft tissue injur*” OR “Back pain” OR “Low back pain” OR “back disorder*” OR “Spinal disease*” OR “Spinal diseases”[MeSH] OR “Back pain”[Mesh] OR “Spinal pain” OR “Sciatica” OR “Sciatic pain” OR “Lumbar radicular pain” OR “Lumbago” OR “Back Ache” OR "Pelvic pain" OR "Pelvic pain"[MeSH] OR “Neck Ache” OR “Neck pain” OR “Arm pain” OR “Shoulder pain” OR “Elbow pain” OR “Hand pain” OR “Wrist pain” OR “Leg pain” OR “Hip pain” OR “Knee pain” OR “Heel pain” OR “Ankle pain” OR ”Foot pain”) | 4,077,642 | 22-02-2023 |
| **#2** | Implementation Strategies | ("Health Plan Implementation"[Mesh] OR "Diffusion of Innovation"[Mesh] OR “Innovation diffusion*” OR "Translational Research, Biomedical"[Mesh] OR "Information Dissemination"[Mesh] OR "Evidence-Based Practice"[Mesh] OR “Implement*” OR “Knowledge transfer*” OR “Knowledge utili*” OR “Knowledge disseminat*” OR “Knowledge adopti*” OR “Knowledge chang*” OR “Knowledge evaluat*” OR “Knowledge use” OR “Knowledge communicat*” OR “Research translat*” OR “Research transfer*” OR “Research utili*” OR “Research disseminat*” OR “Research adopt*” OR “Research chang*” OR “Research evaluat*” OR “Research use” OR “Research communicat*” OR “Evidence translat*” OR “evidence transfer*” OR “evidence utili*” OR “Evidence disseminat*” OR “Evidence adopt*” OR “Evidence chang*” OR “Evidence evaluat*” OR “Evidence use” OR “Evidence communicat*” OR “Translation of knowledge” OR “Translation of research” OR “Translation of evidence” OR “Transfer of knowledge” OR “Transfer of research” OR “Transfer of evidence” OR “Systematic review evidence”) | 821,890 | 22-02-2023 |
| **#3** | Randomised Controlled Trials | ("Randomized Controlled Trial"[Publication Type] OR "Randomized controlled trial"[Title/Abstract] OR “RCT”[Title/Abstract] OR "Randomized controlled stud*"[Title/Abstract] OR “Randomly”[Title/Abstract] OR “Randomized”[Title/Abstract] OR “Randomised”[Title/Abstract]) | 1,227,696 | 22-02-2023 |
| **#1 AND #2** | Musculoskeletal Conditions + Implementation Strategies | (“Musculoskeletal*” OR "Musculoskeletal System"[MeSH] OR "Musculoskeletal Diseases"[Mesh] OR "Musculoskeletal and Neural Physiological Phenomena"[MeSH] OR "Myofascial pain syndromes"[MeSH] OR "Myofascial pain" OR "Musculoskeletal Pain"[MeSH] OR “Arthralgia” OR “Arthralgia”[MeSH] OR “Bursit*” OR “Tendin*” OR “Tendon*” OR “Myalgia” OR “Joint pain” OR “Muscle pain” OR “Soft tissue injur*” OR “Back pain” OR “Low back pain” OR “back disorder*” OR “Spinal disease*” OR “Spinal diseases”[MeSH] OR “Back pain”[Mesh] OR “Spinal pain” OR “Sciatica” OR “Sciatic pain” OR “Lumbar radicular pain” OR “Lumbago” OR “Back Ache” OR "Pelvic pain" OR "Pelvic pain"[MeSH] OR “Neck Ache” OR “Neck pain” OR “Arm pain” OR “Shoulder pain” OR “Elbow pain” OR “Hand pain” OR “Wrist pain” OR “Leg pain” OR “Hip pain” OR “Knee pain” OR “Heel pain” OR “Ankle pain” OR ”Foot pain”) AND ("Health Plan Implementation"[Mesh] OR "Diffusion of Innovation"[Mesh] OR “Innovation diffusion*” OR "Translational Research, Biomedical"[Mesh] OR "Information Dissemination"[Mesh] OR "Evidence-Based Practice"[Mesh] OR “Implement*” OR “Knowledge transfer*” OR “Knowledge utili*” OR “Knowledge disseminat*” OR “Knowledge adopti*” OR “Knowledge chang*” OR “Knowledge evaluat*” OR “Knowledge use” OR “Knowledge communicat*” OR “Research translat*” OR “Research transfer*” OR “Research utili*” OR “Research disseminat*” OR “Research adopt*” OR “Research chang*” OR “Research evaluat*” OR “Research use” OR “Research communicat*” OR “Evidence translat*” OR “evidence transfer*” OR “evidence utili*” OR “Evidence disseminat*” OR “Evidence adopt*” OR “Evidence chang*” OR “Evidence evaluat*” OR “Evidence use” OR “Evidence communicat*” OR “Translation of knowledge” OR “Translation of research” OR “Translation of evidence” OR “Transfer of knowledge” OR “Transfer of research” OR “Transfer of evidence” OR “Systematic review evidence”) | 56,647 | 22-02-2023 |
| **#1 AND #2 AND #3** | Musculoskeletal Conditions + Implementation Strategies + Randomised Controlled Trials | (“Musculoskeletal*” OR "Musculoskeletal System"[MeSH] OR "Musculoskeletal Diseases"[Mesh] OR "Musculoskeletal and Neural Physiological Phenomena"[MeSH] OR "Myofascial pain syndromes"[MeSH] OR "Myofascial pain" OR "Musculoskeletal Pain"[MeSH] OR “Arthralgia” OR “Arthralgia”[MeSH] OR “Bursit*” OR “Tendin*” OR “Tendon*” OR “Myalgia” OR “Joint pain” OR “Muscle pain” OR “Soft tissue injur*” OR “Back pain” OR “Low back pain” OR “back disorder*” OR “Spinal disease*” OR “Spinal diseases”[MeSH] OR “Back pain”[Mesh] OR “Spinal pain” OR “Sciatica” OR “Sciatic pain” OR “Lumbar radicular pain” OR “Lumbago” OR “Back Ache” OR "Pelvic pain" OR "Pelvic pain"[MeSH] OR “Neck Ache” OR “Neck pain” OR “Arm pain” OR “Shoulder pain” OR “Elbow pain” OR “Hand pain” OR “Wrist pain” OR “Leg pain” OR “Hip pain” OR “Knee pain” OR “Heel pain” OR “Ankle pain” OR ”Foot pain”) AND ("Health Plan Implementation"[Mesh] OR "Diffusion of Innovation"[Mesh] OR “Innovation diffusion*” OR "Translational Research, Biomedical"[Mesh] OR "Information Dissemination"[Mesh] OR "Evidence-Based Practice"[Mesh] OR “Implement*” OR “Knowledge transfer*” OR “Knowledge utili*” OR “Knowledge disseminat*” OR “Knowledge adopti*” OR “Knowledge chang*” OR “Knowledge evaluat*” OR “Knowledge use” OR “Knowledge communicat*” OR “Research translat*” OR “Research transfer*” OR “Research utili*” OR “Research disseminat*” OR “Research adopt*” OR “Research chang*” OR “Research evaluat*” OR “Research use” OR “Research communicat*” OR “Evidence translat*” OR “evidence transfer*” OR “evidence utili*” OR “Evidence disseminat*” OR “Evidence adopt*” OR “Evidence chang*” OR “Evidence evaluat*” OR “Evidence use” OR “Evidence communicat*” OR “Translation of knowledge” OR “Translation of research” OR “Translation of evidence” OR “Transfer of knowledge” OR “Transfer of research” OR “Transfer of evidence” OR “Systematic review evidence”) AND ("Randomized Controlled Trial"[Publication Type] OR "Randomized controlled trial"[Title/Abstract] OR “RCT”[Title/Abstract] OR "Randomized controlled stud*"[Title/Abstract] OR “Randomly”[Title/Abstract] OR “Randomized”[Title/Abstract] OR “Randomised”[Title/Abstract]) | 6,698 | 22-02-2023 |

#### Embase

| ***Table A1.3***  *The literature search from Embase* | | | | |
| --- | --- | --- | --- | --- |
| **Search number** | **Search subjects** | **Search** | **Hits** | **Date** |
| **#1** | Musculoskeletal Conditions | (‘Musculoskeletal*’ OR 'Musculoskeletal disease'/exp OR 'Musculoskeletal system'/exp OR 'Musculoskeletal function'/exp OR 'nociceptive pain' OR 'pelvic girdle pain' OR 'pelvic pain'/exp OR 'pelvic pain' OR 'referred pain' OR 'thorax pain' OR 'soft tissue injury'/exp OR ‘soft tissue injury’ OR 'sciatica' OR 'limb pain' OR 'spinal pain' OR 'arthralgia' OR 'bursit*' OR 'tendin*' OR 'tendon*' OR 'myalgia' OR 'joint pain' OR 'muscle pain' OR 'soft tissue injur*' OR 'back pain' OR 'low back pain' OR 'back disorder*' OR 'spinal disease*' OR 'spinal pain' OR 'sciatica' OR 'sciatic pain' OR 'lumbar radicular pain' OR 'lumbago' OR 'back ache' OR 'pelvic pain' OR 'neck ache' OR 'neck pain' OR 'arm pain' OR 'shoulder pain' OR 'elbow pain' OR 'hand pain' OR 'wrist pain' OR 'leg pain' OR 'hip pain' OR 'knee pain' OR 'heel pain' OR 'ankle pain' OR 'foot pain') | 5,093,335 | 22-02-2023 |
| **#2** | Implementation Strategies | (‘Program evaluation’/exp OR ‘Program evaluation’ OR 'Health care planning'/exp OR 'Health care planning' OR 'implementation science' OR ‘Information Dissemination’/exp OR ‘Information Dissemination’ OR 'translational research' OR 'diffusion of innovation' OR ‘Implementation research’ OR 'implementation intention' OR 'health plan implementation' OR 'innovation diffusion*' OR 'evidence-based practice' OR 'implement*' OR 'knowledge transfer*' OR 'knowledge utili*' OR 'knowledge disseminat*' OR 'knowledge adopti*' OR 'knowledge chang*' OR 'knowledge evaluat*' OR 'knowledge use' OR 'knowledge communicat*' OR 'research translat*' OR 'research transfer*' OR 'research utili*' OR 'research disseminat*' OR 'research adopt*' OR 'research chang*' OR 'research evaluat*' OR 'research use' OR 'research communicat*' OR 'evidence translat*' OR 'evidence transfer*' OR 'evidence utili*' OR 'evidence disseminat*' OR 'evidence adopt*' OR 'evidence chang*' OR 'evidence evaluat*' OR 'translation of knowledge' OR 'translation of research' OR 'translation of evidence' OR 'transfer of knowledge' OR 'transfer of research' OR 'transfer of evidence') | 1,330,366 | 22-02-2023 |
| **#3** | Randomised Controlled Trials | ('Randomized controlled trial'/exp OR 'Randomized controlled trial':ti,ab OR ‘Randomized controlled stud*’:ti,ab OR ‘RCT’:ti,ab OR ‘Randomized’:ti,ab OR ‘Randomised’:ti,ab OR ‘Randomly’:ti,ab) | 1,653,137 | 22-02-2023 |
| **#1 AND #2** | Musculoskeletal Conditions + Implementation Strategies | (‘Musculoskeletal*’ OR 'Musculoskeletal disease'/exp OR 'Musculoskeletal system'/exp OR 'Musculoskeletal function'/exp OR 'nociceptive pain' OR 'pelvic girdle pain' OR 'pelvic pain'/exp OR 'pelvic pain' OR 'referred pain' OR 'thorax pain' OR 'soft tissue injury'/exp OR ‘soft tissue injury’ OR 'sciatica' OR 'limb pain' OR 'spinal pain' OR 'arthralgia' OR 'bursit*' OR 'tendin*' OR 'tendon*' OR 'myalgia' OR 'joint pain' OR 'muscle pain' OR 'soft tissue injur*' OR 'back pain' OR 'low back pain' OR 'back disorder*' OR 'spinal disease*' OR 'spinal pain' OR 'sciatica' OR 'sciatic pain' OR 'lumbar radicular pain' OR 'lumbago' OR 'back ache' OR 'pelvic pain' OR 'neck ache' OR 'neck pain' OR 'arm pain' OR 'shoulder pain' OR 'elbow pain' OR 'hand pain' OR 'wrist pain' OR 'leg pain' OR 'hip pain' OR 'knee pain' OR 'heel pain' OR 'ankle pain' OR 'foot pain') AND (‘Program evaluation’/exp OR ‘Program evaluation’ OR 'Health care planning'/exp OR 'Health care planning' OR 'implementation science' OR ‘Information Dissemination’/exp OR ‘Information Dissemination’ OR 'translational research' OR 'diffusion of innovation' OR ‘Implementation research’ OR 'implementation intention' OR 'health plan implementation' OR 'innovation diffusion*' OR 'evidence-based practice' OR 'implement*' OR 'knowledge transfer*' OR 'knowledge utili*' OR 'knowledge disseminat*' OR 'knowledge adopti*' OR 'knowledge chang*' OR 'knowledge evaluat*' OR 'knowledge use' OR 'knowledge communicat*' OR 'research translat*' OR 'research transfer*' OR 'research utili*' OR 'research disseminat*' OR 'research adopt*' OR 'research chang*' OR 'research evaluat*' OR 'research use' OR 'research communicat*' OR 'evidence translat*' OR 'evidence transfer*' OR 'evidence utili*' OR 'evidence disseminat*' OR 'evidence adopt*' OR 'evidence chang*' OR 'evidence evaluat*' OR 'translation of knowledge' OR 'translation of research' OR 'translation of evidence' OR 'transfer of knowledge' OR 'transfer of research' OR 'transfer of evidence') | 108,884 | 22-02-2023 |
| **#1 AND #2 AND #3** | Musculoskeletal Conditions + Implementation Strategies + Randomised Controlled Trials | ('musculoskeletal*' OR 'musculoskeletal disease'/exp OR 'musculoskeletal system'/exp OR 'musculoskeletal function'/exp OR 'nociceptive pain' OR 'pelvic girdle pain' OR 'pelvic pain'/exp OR 'referred pain' OR 'thorax pain' OR 'soft tissue injury'/exp OR 'soft tissue injury' OR 'limb pain' OR 'arthralgia' OR 'bursit*' OR 'tendin*' OR 'tendon*' OR 'myalgia' OR 'joint pain' OR 'muscle pain' OR 'soft tissue injur*' OR 'back pain' OR 'low back pain' OR 'back disorder*' OR 'spinal disease*' OR 'spinal pain' OR 'sciatica' OR 'sciatic pain' OR 'lumbar radicular pain' OR 'lumbago' OR 'back ache' OR 'pelvic pain' OR 'neck ache' OR 'neck pain' OR 'arm pain' OR 'shoulder pain' OR 'elbow pain' OR 'hand pain' OR 'wrist pain' OR 'leg pain' OR 'hip pain' OR 'knee pain' OR 'heel pain' OR 'ankle pain' OR 'foot pain') AND ('program evaluation'/exp OR 'program evaluation' OR 'health care planning'/exp OR 'health care planning' OR 'implementation science' OR 'information dissemination'/exp OR 'information dissemination' OR 'translational research' OR 'diffusion of innovation' OR 'implementation research' OR 'implementation intention' OR 'health plan implementation' OR 'innovation diffusion*' OR 'evidence-based practice' OR 'implement*' OR 'knowledge transfer*' OR 'knowledge utili*' OR 'knowledge disseminat*' OR 'knowledge adopti*' OR 'knowledge chang*' OR 'knowledge evaluat*' OR 'knowledge use' OR 'knowledge communicat*' OR 'research translat*' OR 'research transfer*' OR 'research utili*' OR 'research disseminat*' OR 'research adopt*' OR 'research chang*' OR 'research evaluat*' OR 'research use' OR 'research communicat*' OR 'evidence translat*' OR 'evidence transfer*' OR 'evidence utili*' OR 'evidence disseminat*' OR 'evidence adopt*' OR 'evidence chang*' OR 'evidence evaluat*' OR 'translation of knowledge' OR 'translation of research' OR 'translation of evidence' OR 'transfer of knowledge' OR 'transfer of research' OR 'transfer of evidence') AND ('randomized controlled trial'/exp OR 'randomized controlled trial':ti,ab OR 'randomized controlled stud*':ti,ab OR 'rct':ti,ab OR 'randomized':ti,ab OR 'randomised':ti,ab OR 'randomly':ti,ab) | 10,000 | 22-02-2023 |

#### Scopus

| ***Table A1.4***  *The literature search from Scopus* | | | | |
| --- | --- | --- | --- | --- |
| **Search number** | **Search subjects** | **Search** | **Hits** | **Date** |
| **#1** | Musculoskeletal Conditions | TITLE-ABS-KEY ( ( "musculoskeletal*" OR "myofascial pain" OR "arthralgia" OR "bursit*" OR "tendin*" OR "tendon*" OR "myalgia" OR "joint pain"OR "muscle pain" OR "soft tissue injur*" OR "back pain" OR "low back pain" OR "back disorder*" OR "spinal disease*" OR "spinal pain" OR"sciatica" OR "sciatic pain" OR "lumbar radicular pain" OR "lumbago" OR "back ache" OR "pelvic pain" OR "neck ache" OR "neck pain" OR "arm pain" OR "shoulder pain" OR "elbow pain" OR "hand pain" OR "wrist pain" OR "leg pain" OR "hip pain" OR "knee pain" OR "heel pain" OR "ankle pain" OR "foot pain" ) ) | 705,648 | 22-02-2023 |
| **#2** | Implementation Strategies | TITLE-ABS-KEY ( ( "health plan implementation" OR "diffusion of innovation" OR "innovation diffusion*" OR "translational research, biomedical" OR "information dissemination" OR "evidence-based practice" OR "implement*" OR "knowledge transfer*" OR "knowledge utili*" OR"knowledge disseminat*" OR "knowledge adopti*" OR "knowledge chang*" OR "knowledge evaluat*" OR "knowledge use" OR "knowledge communicat*" OR "research translat*" OR "research transfer*" OR "research utili*" OR "research disseminat*" OR "research adopt*" OR"research chang*" OR "research evaluat*" OR "research use" OR "research communicat*" OR "evidence translat*" OR "evidence transfer*" OR"evidence utili*" OR "evidence disseminat*" OR "evidence adopt*" OR "evidence chang*" OR "evidence evaluat*" OR "evidence use" OR"evidence communicat*" OR "translation of knowledge" OR "translation of research" OR "translation of evidence" OR "transfer of knowledge"OR "transfer of research" OR "transfer of evidence" OR "systematic review evidence" ) ) | 3,521,271 | 22-02-2023 |
| **#3** | Randomised Controlled Trials | TITLE-ABS-KEY ("Randomized Controlled Trial" OR "Randomized controlled trial" OR “RCT” OR "Randomized controlled stud*" OR “Randomly” OR “Randomized” OR “Randomised”) | 1,793,015 | 22-02-2023 |
| **#1 AND #2** | Musculoskeletal Conditions + Implementation Strategies | ( TITLE-ABS-KEY ( ( "musculoskeletal*" OR "myofascial pain" OR "arthralgia" OR "bursit*" OR "tendin*" OR "tendon*" OR "myalgia" OR "joint pain"OR "muscle pain" OR "soft tissue injur*" OR "back pain" OR "low back pain" OR "back disorder*" OR "spinal disease*" OR "spinal pain" OR "sciatica"OR "sciatic pain" OR "lumbar radicular pain" OR "lumbago" OR "back ache" OR "pelvic pain" OR "neck ache" OR "neck pain" OR "arm pain" OR"shoulder pain" OR "elbow pain" OR "hand pain" OR "wrist pain" OR "leg pain" OR "hip pain" OR "knee pain" OR "heel pain" OR "ankle pain" OR"foot pain" ) ) AND TITLE-ABS-KEY ( ( "health plan implementation" OR "diffusion of innovation" OR "innovation diffusion*" OR "translational research, biomedical" OR "information dissemination" OR "evidence-based practice" OR "implement*" OR "knowledge transfer*" OR "knowledge utili*" OR "knowledge disseminat*" OR "knowledge adopti*" OR "knowledge chang*" OR "knowledge evaluat*" OR "knowledge use" OR"knowledge communicat*" OR "research translat*" OR "research transfer*" OR "research utili*" OR "research disseminat*" OR "research adopt*"OR "research chang*" OR "research evaluat*" OR "research use" OR "research communicat*" OR "evidence translat*" OR "evidence transfer*" OR"evidence utili*" OR "evidence disseminat*" OR "evidence adopt*" OR "evidence chang*" OR "evidence evaluat*" OR "evidence use" OR "evidence communicat*" OR "translation of knowledge" OR "translation of research" OR "translation of evidence" OR "transfer of knowledge" OR "transfer of research" OR "transfer of evidence" OR "systematic review evidence" ) ) ) | 15,645 | 22-02-2023 |
| **#1 AND #2 AND #3** | Musculoskeletal Conditions + Implementation Strategies + Randomised Controlled Trials | TITLE-ABS-KEY ( "musculoskeletal*" OR "myofascial pain" OR "arthralgia" OR "bursit*" OR "tendin*" OR "tendon*" OR "myalgia" OR "joint pain" OR"muscle pain" OR "soft tissue injur*" OR "back pain" OR "low back pain" OR "back disorder*" OR "spinal disease*" OR "spinal pain" OR "sciatica" OR"sciatic pain" OR "lumbar radicular pain" OR "lumbago" OR "back ache" OR "pelvic pain" OR "neck ache" OR "neck pain" OR "arm pain" OR "shoulder pain" OR "elbow pain" OR "hand pain" OR "wrist pain" OR "leg pain" OR "hip pain" OR "knee pain" OR "heel pain" OR "ankle pain" OR "foot pain" )AND TITLE-ABS-KEY ( "health plan implementation" OR "diffusion of innovation" OR "innovation diffusion*" OR "translational research, biomedical"OR "information dissemination" OR "evidence-based practice" OR "implement*" OR "knowledge transfer*" OR "knowledge utili*" OR "knowledge disseminat*" OR "knowledge adopti*" OR "knowledge chang*" OR "knowledge evaluat*" OR "knowledge use" OR "knowledge communicat*" OR"research translat*" OR "research transfer*" OR "research utili*" OR "research disseminat*" OR "research adopt*" OR "research chang*" OR "research evaluat*" OR "research use" OR "research communicat*" OR "evidence translat*" OR "evidence transfer*" OR "evidence utili*" OR "evidence disseminat*" OR "evidence adopt*" OR "evidence chang*" OR "evidence evaluat*" OR "evidence use" OR "evidence communicat*" OR "translation of knowledge" OR "translation of research" OR "translation of evidence" OR "transfer of knowledge" OR "transfer of research" OR "transfer of evidence"OR "systematic review evidence" ) AND TITLE-ABS-KEY ( "randomized controlled trial" OR "randomized controlled trial" OR "rct" OR "randomized controlled stud*" OR "randomly" OR "randomized" OR "randomised" ) | 2,035 | 22-02-2023 |

#### Cochrane (CENTRAL)

| ***Table A1.5***  *The literature search from Cochrane (CENTRAL)* | | | | |
| --- | --- | --- | --- | --- |
| **Search number** | **Search subjects** | **Search** | **Hits** | **Date** |
| **#1** | Musculoskeletal Conditions | (“Musculoskeletal*” OR [mh "Musculoskeletal Diseases"] OR [mh "Musculoskeletal and Neural Physiological Phenomena"] OR [mh "Myofascial pain syndromes"] OR "Myofascial pain" OR [mh “Musculoskeletal Pain”] OR “Arthralgia” OR [mh “Arthralgia”] OR “Bursit*” OR “Tendin*” OR “Tendon*” OR “Myalgia” OR “Joint pain” OR “Muscle pain” OR “Soft tissue injur*” OR “Back pain” OR “Low back pain” OR “back disorder*” OR “Spinal disease*” OR [mh “Spinal diseases”] OR [mh “Back pain”] OR “Spinal pain” OR “Sciatica” OR “Sciatic pain” OR “Lumbar radicular pain” OR “Lumbago” OR “Back Ache” OR "Pelvic pain" OR “Neck Ache” OR “Neck pain” OR “Arm pain” OR “Shoulder pain” OR “Elbow pain” OR “Hand pain” OR “Wrist pain” OR “Leg pain” OR “Hip pain” OR “Knee pain” OR “Heel pain” OR “Ankle pain” OR ”Foot pain”) | 199,517 | 22-02-2023 |
| **#2** | Implementation Strategies | ([mh "Health Plan Implementation"] OR [mh "Diffusion of Innovation"] OR “Innovation diffusion*” OR [mh "Translational Research, Biomedical"] OR [mh "Information Dissemination"] OR [mh "Evidence-Based Practice"] OR [mh "Implementation Science"] OR [mh "Implementation Plan, Annual"] OR “Implement*” OR “Knowledge transfer*” OR “Knowledge utili*” OR “Knowledge disseminat*” OR “Knowledge adopti*” OR “Knowledge chang*” OR “Knowledge evaluat*” OR “Knowledge use” OR “Knowledge communicat*” OR “Research translat*” OR “Research transfer*” OR “Research utili*” OR “Research disseminat*” OR “Research adopt*” OR “Research chang*” OR “Research evaluat*” OR “Research use” OR “Research communicat*” OR “Evidence translat*” OR “evidence transfer*” OR “evidence utili*” OR “Evidence disseminat*” OR “Evidence adopt*” OR “Evidence chang*” OR “Evidence evaluat*” OR “Evidence use” OR “Evidence communicat*” OR “Translation of knowledge” OR “Translation of research” OR “Translation of evidence” OR “Transfer of knowledge” OR “Transfer of research” OR “Transfer of evidence” OR “Systematic review evidence”) | 14,753 | 22-02-2023 |
| **#3** | Randomised Controlled Trials | **Randomized Controlled Trial**  ([mh “Randomized Controlled Trial”] OR "Randomized Controlled Trial":ti,ab,kw OR "Randomized controlled trial":ti,ab,kw OR “RCT”:ti,ab,kw OR "Randomized controlled stud*":ti,ab,kw OR “Randomly”:ti,ab,kw OR “Randomized”:ti,ab,kw OR “Randomised”:ti,ab,kw) | 1,164,840 | 22-02-2023 |
| **#1 AND #2** | Musculoskeletal Conditions + Implementation Strategies | (“Musculoskeletal*” OR [mh "Musculoskeletal Diseases"] OR [mh "Musculoskeletal and Neural Physiological Phenomena"] OR [mh "Myofascial pain syndromes"] OR "Myofascial pain" OR [mh “Musculoskeletal Pain”] OR “Arthralgia” OR [mh “Arthralgia”] OR “Bursit*” OR “Tendin*” OR “Tendon*” OR “Myalgia” OR “Joint pain” OR “Muscle pain” OR “Soft tissue injur*” OR “Back pain” OR “Low back pain” OR “back disorder*” OR “Spinal disease*” OR [mh “Spinal diseases”] OR [mh “Back pain”] OR “Spinal pain” OR “Sciatica” OR “Sciatic pain” OR “Lumbar radicular pain” OR “Lumbago” OR “Back Ache” OR "Pelvic pain" OR “Neck Ache” OR “Neck pain” OR “Arm pain” OR “Shoulder pain” OR “Elbow pain” OR “Hand pain” OR “Wrist pain” OR “Leg pain” OR “Hip pain” OR “Knee pain” OR “Heel pain” OR “Ankle pain” OR ”Foot pain”) AND ([mh "Health Plan Implementation"] OR [mh "Diffusion of Innovation"] OR “Innovation diffusion*” OR [mh "Translational Research, Biomedical"] OR [mh "Information Dissemination"] OR [mh "Evidence-Based Practice"] OR [mh "Implementation Science"] OR [mh "Implementation Plan, Annual"] OR “Implement*” OR “Knowledge transfer*” OR “Knowledge utili*” OR “Knowledge disseminat*” OR “Knowledge adopti*” OR “Knowledge chang*” OR “Knowledge evaluat*” OR “Knowledge use” OR “Knowledge communicat*” OR “Research translat*” OR “Research transfer*” OR “Research utili*” OR “Research disseminat*” OR “Research adopt*” OR “Research chang*” OR “Research evaluat*” OR “Research use” OR “Research communicat*” OR “Evidence translat*” OR “evidence transfer*” OR “evidence utili*” OR “Evidence disseminat*” OR “Evidence adopt*” OR “Evidence chang*” OR “Evidence evaluat*” OR “Evidence use” OR “Evidence communicat*” OR “Translation of knowledge” OR “Translation of research” OR “Translation of evidence” OR “Transfer of knowledge” OR “Transfer of research” OR “Transfer of evidence” OR “Systematic review evidence”) | 1,538 | 22-02-2023 |
| **#1 AND #2 AND TRIALS** | Musculoskeletal Conditions + Implementation Strategies + Filter: Trials | (“Musculoskeletal*” OR [mh "Musculoskeletal Diseases"] OR [mh "Musculoskeletal and Neural Physiological Phenomena"] OR [mh "Myofascial pain syndromes"] OR "Myofascial pain" OR [mh “Musculoskeletal Pain”] OR “Arthralgia” OR [mh “Arthralgia”] OR “Bursit*” OR “Tendin*” OR “Tendon*” OR “Myalgia” OR “Joint pain” OR “Muscle pain” OR “Soft tissue injur*” OR “Back pain” OR “Low back pain” OR “back disorder*” OR “Spinal disease*” OR [mh “Spinal diseases”] OR [mh “Back pain”] OR “Spinal pain” OR “Sciatica” OR “Sciatic pain” OR “Lumbar radicular pain” OR “Lumbago” OR “Back Ache” OR "Pelvic pain" OR “Neck Ache” OR “Neck pain” OR “Arm pain” OR “Shoulder pain” OR “Elbow pain” OR “Hand pain” OR “Wrist pain” OR “Leg pain” OR “Hip pain” OR “Knee pain” OR “Heel pain” OR “Ankle pain” OR ”Foot pain”) AND ([mh "Health Plan Implementation"] OR [mh "Diffusion of Innovation"] OR “Innovation diffusion*” OR [mh "Translational Research, Biomedical"] OR [mh "Information Dissemination"] OR [mh "Evidence-Based Practice"] OR [mh "Implementation Science"] OR [mh "Implementation Plan, Annual"] OR “Implement*” OR “Knowledge transfer*” OR “Knowledge utili*” OR “Knowledge disseminat*” OR “Knowledge adopti*” OR “Knowledge chang*” OR “Knowledge evaluat*” OR “Knowledge use” OR “Knowledge communicat*” OR “Research translat*” OR “Research transfer*” OR “Research utili*” OR “Research disseminat*” OR “Research adopt*” OR “Research chang*” OR “Research evaluat*” OR “Research use” OR “Research communicat*” OR “Evidence translat*” OR “evidence transfer*” OR “evidence utili*” OR “Evidence disseminat*” OR “Evidence adopt*” OR “Evidence chang*” OR “Evidence evaluat*” OR “Evidence use” OR “Evidence communicat*” OR “Translation of knowledge” OR “Translation of research” OR “Translation of evidence” OR “Transfer of knowledge” OR “Transfer of research” OR “Transfer of evidence” OR “Systematic review evidence”)  **Filter: TRIALS** | 1,210 | 22-02-2023 |
