## Supplementary material for "Effectiveness of Implementation Interventions in Musculoskeletal Healthcare: A Systematic Review": Definition of outcome domains

### Additional file 3 - Development & Definitions of Outcome Domains

#### Development of Outcome Domains

Outcome domains for HCP outcomes were established by a two-round consensus discussion process. In the first round, all primary outcome measures from the included studies were extracted into preliminary domains and frequency across the included studies were established. In the second round, broader domains for outcomes emerged, and encompassed the group of previously established domains (***Figure A2.1***).

***Figure A2.1***

*Overview of outcome domains in the included studies (left) and the condensed outcome concept domains (right).*

#### Definitions of Outcome Concept Domains

We pragmatically mapped and formulated outcome domains for HCP’s in the absence of an established core outcome set. However, inspiration for these HCP outcome domains was drawn from Proctor et al. 2011. We specifically aimed for outcome concepts, such as fidelity and adoption, as described by Proctor et al. 2011. However, we decided to term them “adherence to implemented intervention” and “uptake of knowledge”, as the majority of the included literature in this review used the terms “adherence” and “uptake”, which are in alignment with Proctor et al. 2011, who also recognize that these terms are frequently used in the literature.

Due to the varied ways of measuring "adherence" in the included studies, we argue that some of them were measuring uptake rather than adherence. This distinction was evident in studies that implemented guidelines which dictated recommended practice behaviour. However, in some studies, adherence was then measured using questionnaires related to hypothetical scenarios (vignettes). While this may indicate the clinician's intention, it cannot be assumed that they would act the same way in a real clinical situation. Consequently, these outcomes were categorised as "Uptake of knowledge" rather than "Adherence to implemented intervention," even though they were referred to as "adherence to guideline" in some studies. Given these differences, we deemed it necessary to provide specific definitions for each domain. See definitions of outcomes in ***Table A3.1***.

| ***Table A3.1***  *Definition of Outcome concept domains* | |
| --- | --- |
| **Outcome concept domains** | **Definitions** |
| **Definition of outcome concept domains for HCPs** | |
| Adherence to implemented intervention | Any measurement regarding practice behaviour is concordant with the implemented intervention, such as a guideline or a product. Either by assessing the patient records, auditing files, or by having an assessor observe the clinician, either in real time, or by video recording. The word “intervention” is used in place of the implemented product. |
| Uptake of knowledge | Any measurement that assesses how well the clinician has understood and remembered the implemented product, often assessed with questionnaires, multiple choice tests, and work on vignettes. |
| Referral to imaging | Any measurement that assesses the impact from implementation interventions on referrals to medical imaging investigations, such as (but not limited to) x-ray, CT-scan, and MRI-scans. |
| Referral to secondary care | Any measurement that assesses the impact on referrals to secondary care, such as (but not limited to) physiotherapy, chiropractic, specialist’s doctors, podiatrists, dieticians etc. |
| Prescription of analgesics | Any measurement that assesses the impact on prescriptions to any type of medicine that is considered an analgetic painkiller. |
| Clinician satisfaction | Any measurement assessing the level of satisfaction the clinician had with the implementation process |
| **Definition of outcome concept domains for patients** | |
| Function | Any measurement assessing the level of functioning on patient level due to musculoskeletal disorders. Could involve PRO’s (Such as ODI, RMDQ, NDI, PSFS, PROMIS-PF20 and the like) |
| Pain | Any measurement assessing the pain impact on the patient, such as NPRS, VAS, 3 items pain means and the like due to musculoskeletal conditions. |
| Workability | Any measurement assessing the impact on the patient’s ability to maintain their occupation, such as sick leave and sick days due to musculoskeletal disorders. |
| Physical activity | Any measurement assessing the amount of physical activity the patients perform after an intervention. |
| Patient satisfaction | Any measurement assessing the level of satisfaction patients have with the received treatment or received outcome. |
| Quality of life | Any measurement assessing the quality of life with PROs such as EQ-5D or SF-36. |
| Patient adherence | Any measurements assessing the level of adherence the patient have to the treatment. |
| **Definition of outcome concept domains for implementation costs** | |
| Cost-effectiveness | Any measurement of direct and indirect cost for the patients or any analysis looking at cost-effectiveness including cost-utility depending on two values such as health related quality of life and cost. |
| **Abbreviations:** CT: Computerised tomography; MRI: Magnetic resonance imaging; PRO: Patient reported outcomes; ODI: Oswestry disability index; RMDQ: Roland-Morris disability questionnaire; NDI: Neck disability index; PSFS: Patient specific functional scale; PROMIS-PF20: Patient reported outcome measurement information system – Physical function-20; NPRS: Numeric pain rating scale; VAS: Visual analogue scale; EQ-5D: European quality of life – 5 Dimensions; SF-36: Short form - 36 | |
