## Supplementary material for "Effectiveness of Implementation Interventions in Musculoskeletal Healthcare: A Systematic Review": Full version of table 1

### Additional file 4 – Full Version of Table 1

| ***Table A4.1***  *Full Version of Table 1 - Study Characteristics of All Included Studies* | | | | | |
| --- | --- | --- | --- | --- | --- |
| **Author,, Year**  **Study Design**  **Country of Origin** | **HCP’s (n)**  **Patient condition (n)**  **Clinical setting (n)** | **Implementation clusters & strategies** | | **Primary Outcomes**  *Healthcare Professionals; Patients; Cost-effectiveness* | **Findings** |
|  |  | *Implementation strategy and Waltz’s Implementation Clusters* | *Comparator* |  |  |
| Becker et al., 2008  Cluster RCT  Germany | GP’s (126)  Patients with LBP (1378)  Primary care (118 practices) | **Guideline Implementation (GI) group**  **Description of Implementation Strategy:**  GPs were trained in using the LBP guideline of the German College of General Practitioners and Family Physicians (DEGAM).  Three interactive seminars for participating GPs were held.  The seminars included information, education and discussions.  Individual educational visits by study nurses to participating GPs twice to hand over the guideline and discuss individual problems with GI.  **Waltz’s Classification:**  *Develop stakeholder interrelationships.*   - Conduct local consensus discussions.   *Train and educate stakeholders.*   - Conduct ongoing training. - Provide ongoing consultation. - Make training dynamic. - Distribute educational materials. - Conduct educational meetings. - Conduct educational outreach visits.   **Guideline Implementation (GI) + Motivational Counselling (MC) group**  **Description of Implementation Strategy:**  In addition to the GI group the GI + MC group were introduced to MC strategies during the third educational session and two nurses per practice received a 20-hour training (2 full-day workshops and 1–3 supervision sessions).  Practice nurses were contacted regularly by study coordinators to identify barriers and problems.  **Waltz’s Classification:**  In addition to the GI group, the GI + MC group received:  *Use evaluative and iterative strategies.*   - Assess for readiness and identify barriers and facilitators.   *Provide interactive assistance.*   - Provide clinical supervision | **Description of implementation Strategy:**  The control group received the guideline via mail.  **Waltz’s classification:**  *Train and educate stakeholders.*   - Distribute educational materials. | **Patients:**  *Function*  Functional capacity by 12-item Hannover Functional Ability Questionnaire for Measuring Back Pain Related Functional Limitations | **6 months follow-up:**   - The GI group did not score higher on functional capacity than the control group with a mean difference of 2.652 (95% CI: -0.704 to 6.007, p=0.120) - The GI+MC group scored higher on functional capacity than the control group, with a mean difference of 3.650 (95% CI: 0.320 to 6.979, p=0.032) |
| Becker et al., 2012 Cost-effectiveness analysis of a cluster RCT Germany | GP’s (126)  Patients with LBP (1322)  Primary care (118 practices) | This cost-effectiveness study was performed using data from Becker et al., 2008.  For a description of the implementation strategies and Waltz’s Classification, see Becker et al., 2008. | | **Cost-Effectiveness:**  Total costs   - Direct costs - Indirect costs | **1-6 months follow-up:**  For Indirect cost the GI and GI+MC group had lower costs than the control group, with a mean difference of -332.51€ (95%CI: -650 to -44) and -302.83€ (95% CI: -621 to -6).  For Total cost the GI+MC group had lower costs than the control group, with a mean difference of -482.59€ (95% CI: −983 to -54). |
| Bekkering et al., 2005 (a)  Cluster RCT  The Netherlands | Physiotherapists (113)  Patients with LBP (500)  Primary care  (68 practices) | **Description of Implementation Strategy:**  Received the guideline by mail, along with four forms to self-evaluate and facilitate discussion. In addition, an article concerning the guidelines was published.  They also received two training sessions, each lasting 2.5 hours, along with 2 hours preparation time for each session.  The sessions included interventions, such as expert advice, interactive education like role-plays with an actor, discussion, feedback and reminders.  **Waltz’s Classification:**  *Provide interactive assistance.*   - Provide clinical supervision.   *Develop stakeholder interrelationships*.   - Conduct local consensus discussions.   *Train and educate stakeholders*.   - Provide ongoing consultation. - Make training dynamic. - Distribute educational materials. - Conduct educational meetings. - Conduct educational outreach visits.   *Support clinicians*   - Remind clinicians | **Description of Implementation Strategy:**  Received the guideline by mail, along with four forms to self-evaluate and facilitate discussion.  In addition, an article concerning the guidelines was published.  **Waltz’s Classification:**  *Train and educate stakeholders.*   - Distribute educational materials | **Healthcare Professionals:**  *Adherence to implemented intervention.*   - Limit number of sessions - Use functional treatment goals. - Use active interventions.   Give adequate information | Physiotherapists in the intervention group, compared to the control group:   - Correctly limited number of treatment sessions with an OR of 2.39 (95% CI: 1.12 to 5.12) - More frequent functional goalsetting with an OR of 1.99 (95% CI: 1.06 to 3.72) - Frequency of active interventions with an OR of 2.79 (95% CI: 1.19 to 6.55) - Gave adequate advice with an OR of 3.59 (95% CI: 1.35 to 9.55)   All four recommendations with an OR 2.05 (95% CI: 1.15 to 3.65) |
| Bekkering et al., 2005 (b)  Cluster RCT  The Netherlands | Physiotherapists (113)  Patients with LBP (500)  Primary care (68 practices) | This study was performed using data from Bekkering et al., 2005 (a).  For a description of the implementation strategies and Waltz’s Classification, see Bekkering et al., 2005 (a). | | **Patients:**  *Function*   - Physical functioning (the Quebec Back Pain Disability Scale [QBPDS])   *Pain*   - Pain intensity (NRS)   *Workability*   - Sick leave (Number of days off work in the last 6 weeks) | **12 weeks follow-up:**  The intervention group did not score higher in physical functioning than the control group with a mean difference of 2.83 points on the QBPDS (95% CI: -0.66 to 6.31).  The intervention group did not score higher in physical functioning than the control group with a mean difference of 0.34 (95% CI: -0.19 to 0.88).  No analyses were performed for sick leave.  **52 weeks/12 months follow-up:**  At 12 months there was no difference between the 2 groups on physical functioning (X^2^=4.88, df=4, p>0.05) or on pain (X^2^=6.05, df=4, p>0.05). |
| Bishop et al., 2006  RCT  Canada | Family physicians (462)  Patients with LBP (428)  Primary care. | **Group 2:**  **Description of Implementation Strategy:**  In Group 2, the patient’s family physician received a copy of the clinical practice guidelines.  In addition, each family physician received a ‘‘guideline reminder letter’’ at each of three separate stages of the patient’s clinical course summarizing the different aspects of the guidelines that specifically applied to the 0–4-week, 5–12-week, and greater than 12-week postinjury periods.  **Waltz’s Classification:**  *Train and educate stakeholders.*   - Distribute educational materials.   *Support clinicians*   - Remind clinicians.   **Group 3:**  **Description of Implementation Strategy:**  In addition to intervention in Group 2 the family physician in Group 3 the individual patient received lay language versions of a pamphlet outlining the clinical practice guidelines.  **Waltz’s Classification:**  In addition to Group 2, Group 3 received:  *Engage consumers.*   - Involve patients/consumers and family members | **Group 1:**  No intervention | **Healthcare Professionals:**  *Adherence to implemented intervention.*   - Guideline concordant patient assessment (History, past history) and examination (Neurological testing, regional back examination, red flag screening) - Guideline treatment recommendation, separated into a) Concordant, or b) Discordant. Each category had specific criteria at 3 time points (0-4 weeks, 5-12 weeks, >12 weeks) - Concordant treatment: Education and reassurance, aerobic exercise, non-narcotic medication, physical therapy modalities, spinal manipulation withing the 4^th^ week. Bed rest not greater than 4 days. Work conditioning programs within the 5–12-week time period. Return to full or modified work at 12 weeks- - Discordant treatment: Routine use of narcotic analgesics, more than 4 days of bed rest, Physical therapy modalities after the 4^th^ week, Continued use passive therapy of spinal manipulative therapy; recycling through treatments and programs that had previously failed after 12 weeks | **Recorded history and physical findings:**  Group 2 compared to control: p=0.89  Group 3 compared to control: p=0.90  **Concordance weeks 0-4 (Medication)**  Group 2 compared to control: p=0.14  Group 3 compared to control: p=0.08  **Concordance weeks 0-4 (recommended exercise)**  Group 3 compared to control: p=0.05  **Concordance weeks 5-12 (Supervised exercise)**  Group 2 compared to control: p=0.11  Group 3 compared to control: p=0.18  **Concordance weeks > 12 weeks (Return to work)**  Group 2 compared to control: p=0.07  Group 3 compared to control: p=0.14  **Discordance weeks 0-4 (Extended bed rest)**  Group 2 compared to control: p=0.05  **Discordance weeks 5-12 (Continued use of passive therapies)**  Group 2 compared to control: p=0.04  Group 3 compared to control: p=0.05 |
| Bruyndonckx et al., 2018  Cluster RCT Belgium | GP’s (4530)  Patients with LBP  Primary care (3401) practices) | **Description of Implementation Strategy:**  The academic detailer visited the clinics for 15-20 minutes and provided four comprehensible key messages about prescriptions of analgesics for patients with osteoarthritis.  **Waltz’s Classification:**  *Train and educate stakeholders*   - Conduct educational outreach visits | No intervention. | **Healthcare Professionals:**  *Prescription of analgesics*  Each key message was evaluated by:   - Odds of being reimbursed for an analgesic or NSAID. - Average defined daily dose of paracetamol per patient reimbursed for paracetamol per month. - Odds of being reimbursed for a recommended NSAID when reimbursed for any NSAID. - Odds of being reimbursed for an NSAID and a PPI when reimbursed for an NSAID. | When comparing the intervention group to the control group, the data suggested:   - Odds of being reimbursed for an analgesic or NSAID: β6 0.9980 (99% CI: 0.9834 to 1.0128) β7 0.9889 )99% CI: 0.9767 to 1.0013) - Average defined daily dose of paracetamol per patient reimbursed for paracetamol per month: β6 0.3287 (99% CI: − 1.0491 to 0.3917) β7 0.0386 (99% CI: − 0.1973 to 0.2745) - Odds of being reimbursed for a recommended NSAID when reimbursed for any NSAID: β6 1.1903 (99% CI: 1.0757 to 1.3171) p<0.01 β7 0.9901 (99% CI: 0.9719 to 1.0086) - Odds of being reimbursed for an NSAID and a PPI when reimbursed for an NSAID: β6 1.0217 (99% CI: 0.9243 to 1.1294) - β7 0.9926 (99% CI: 0.9685 to 1.0173) |
| Bussières et al., 2010  RCT  Switzerland | Chiropractors (160)  Patients with LBP  Private practice | All participants in both the experimental and control groups first attended a 20-minute lecture entitled Diagnostic Imaging Practice Guidelines for Musculoskeletal Complaints in Adults: An Evidence Based Approach.  **Intervention group 1 (IG1)**  **Description of Implementation Strategy:**  A 90-minute educational workshop was presented to the intervention group by two chiropractic specialists. Topics covered included evidence-based recommendations contained in diagnostic imaging guidelines for spine disorders and this was underpinned with 10 case scenarios.  **Waltz’s Classification:**  *Develop stakeholder interrelationships.*   - Conduct local consensus discussions.   *Train and educate stakeholders.*   - Make training dynamic. - Conduct educational meetings.   **Intervention group 2 (IG2)**  **Description of Implementation Strategy:**  In addition to the educational workshop IG2 was invited to review the PowerPoint presented at the educational workshop 6–8 weeks after the conference, acting as a reminder at midpoint.  **Waltz’s Classification:**  In addition to the educational workshop IG2 received:  *Train and educate stakeholders.*   - Conduct ongoing training.   *Support clinicians*   - Remind clinicians.   **Intervention group 3 (IG3)**  **Description of Implementation Strategy:**  IG3 received no invitation to review the PowerPoint presented at the educational workshop or reminder at midpoint and were not informed of this reminder and additional intervention.  **Waltz’s Classification:**  IG3 received no additional interventions than IG1. | **Description of Implementation Strategy:**  The control group attended a chiropractic technique seminar on spinal pain where no discussion on the use of imaging took place.  **Waltz’s Classification:**  *Develop stakeholder interrelationships.*   - Conduct local consensus discussions.   *Train and educate stakeholders.*   - Make training dynamic. - Conduct educational meetings | **Healthcare Professionals:**  *Uptake of knowledge*  Rate of appropriate responses for the use of diagnostic imaging through three questionnaires - each consisting of 10 different spine case scenarios. | IG2 increased at the post-test by 16.4% compared to baseline with a mean change (SD) of 0.785 (1.53), which was significantly greater than the comparison group (IG1) with a mean change (SD) of 0.647 (2.06) for the same period (p = 0.043).  Scores for the pretest and the final test for all four groups were not significantly different (p = 0.348). |
| Chipchase et al., 2016  RCT  Australia | Physiotherapists (23)  Patients with neck pain (158) Physiotherapy Practice | **Description of implementation Strategy:**  A two-day workshop provided an evidence-based approach towards the diagnosis and management of neck disorders, with an emphasis on multimodal interventions inclusive of advice, education, exercise and manual therapy.  The workshop consisted of lectures demonstrations, practice, supervision and discussion over two-days.  A five-hour follow-up session were given one month later providing participants with the opportunity for further skills practice, reflection and discussion.  **Waltz’s Classification:**  *Provide interactive assistance.*   - Provide clinical supervision.   *Develop stakeholder interrelationships.*   - Conduct local consensus discussions.   *Train and educate stakeholders.*   - Conduct ongoing training. - Provide ongoing consultation. - Make training dynamic. - Conduct educational meetings | **Description of implementation Strategy:**  The control received the same two-day workshop but did not receive a follow up session.  **Waltz’s Classification:**  *Provide interactive assistance.*   - Provide clinical supervision.   *Develop stakeholder interrelationships.*   - Conduct local consensus discussions.   *Train and educate stakeholders.*   - Conduct ongoing training. - Provide ongoing consultation. - Make training dynamic. - Conduct educational meetings | **Healthcare Professionals:**  *Uptake of knowledge*  A questionnaire measuring ‘practice behavior’ with two sections:   - Section 1 concerning confidence in the assessment of cervical motor and sensorimotor function and the prescription and progression of exercise. - Section 2 concerning usual management strategies.   **Patients:**  *Function*   - Neck Disability Index (NDI) | Both groups significantly improved in all areas on ‘practice behavior’ from baseline, but no significant between-group differences were identified for any of the Likert scale responses in section 1.  No significant between-group differences were Following the educational intervention, there was no change in practitioners' reports of the frequency with which they used any of these management strategies (p>0.05).  There were no differences between groups in terms of patient outcomes as measured by the NDI before and after the educational interventions (F= 2.88, df=1, p=0.11, n^2^=0.03 [95% CI: -0.04 to 0.10]). |
| Cleland et al., 2009  RCT  USA | Physiotherapists (30)  Patients with neck pain (1199)  Physiotherapy practices | **Description of implementation Strategy:**  All physiotherapists attended a 2-day (4-hours a day) continuous education course on the management of neck pain, presented by specialist physiotherapists. It included approximately 25% lectures, and 75% practical sessions. Additionally had two 1.5-hour educational meetings with the same specialist physiotherapists 4 and 7 weeks are the course. Further an individual outreach visits with a 1-hour co-treatment of the participants patient.  **Waltz’s Classification:**  *Provide interactive assistance.*   - Provide clinical supervision.   *Train and educate stakeholders.*   - Conduct ongoing training. - Provide ongoing consultation. - Conduct educational meetings. - Conduct educational outreach visits | **Description of implementation Strategy:**  Attended the continuous education course but did not receive any follow-up.  **Waltz’s Classification:**  *Train and educate stakeholders.*   - Conduct educational meetings. | **Patients:**  *Function  -* Neck disability index (NDI)  *Pain*  - Numeric pain rating scale (NPRS) | Post-test data showed the following.  *Function:*  MD 3.7 (95% CI: 0.84 to 6.5) p=0.013 between groups in favour of the intervention group  *Pain:*  MD 0.38 (95% CI: -0.057 to 0.82) p=0.088 between groups in favour of the intervention group |
| Coombs et al., 2021 Stepped wedge cluster RCT Australia. | Rheumatologists, physiotherapists and emergency physicians (269)  Patients with LBP (4491)  Primary care | **Description of Implementation Strategy:**  Five main components (for elaboration see article) consisting of education seminars, development and distribution of educational material, provision of non-opioid pain management, giving access to fast-track referrals to outpatient services, and audit and feedback.  **Waltz’s Classification:**  *Use evaluative and iterative strategies.*   - Assess for readiness and identify barriers and facilitators. - Audit and provide feedback.   *Adapt and tailor to context.*   - Tailor strategies   *Train and educate stakeholders.*   - Develop educational materials. - Distribute educational materials. - Conduct educational outreach visits | No intervention | **Healthcare Professionals:**  *Referral to imaging*   - Proportion of patients receiving Lumbar imaging | The intervention did not significantly reduce the odds of lumbar imaging OR=0.77 (95% CI of 0.47 to 1.26) |
| Dey et al., 2004 Cluster RCT England | GP’s  Patients with LBP (2187)  Primary care (24 health centres) | **Description of implementation strategy:**  Two members of the implementation team attended each meeting and facilitated structured interactive discussion. Posters were developed and given to the GP’s reinforcing the guideline recommendations.  **Waltz’s classification:**  *Use evaluative and iterative strategies.*   - Assess for readiness and identify barriers and facilitators.   *Provide interactive assistance.*   - Facilitation   *Adapt and tailor to context.*   - Tailor strategies   *Develop stakeholder interrelationships.*   - Conduct local consensus discussions.   *Train and educate stakeholders.*   - Distribute educational materials. - Conduct educational outreach visits | No intervention | **Healthcare Professionals:**  *Adherence to implemented intervention.*   - Issued a sickness certificate.   *Referral to imaging*   - Referral to x-ray   *Referral to secondary care*   - Referred to secondary care.   *Prescription of analgesics*   - Prescribed opioids or muscle relaxants | - Issued a sickness certificate: X^2^ = 0.11; 1 degrees of freedom; p=0.74 - Referral to x-ray: X^2^ = 0.24; 1 degrees of freedom; p=0.62 - Referred to secondary care: X^2^ = 2.36; 1 degrees of freedom; p=0.12 - Prescribed opioids or muscle relaxants: X^2^ = 0.00014; 1 degrees of freedom; p=0.99 |
| Eccles et al., 2001  Cluster RCT  England and Scotland | GPs (162)  Patients with LBP and knee pain (1693)  Primary care (48 practices) | **Audit & feedback group:** **Description of Implementation Strategy:**  The referral guidelines were sent by post to all GPs.  Feedback (prepared by the research team from routine data provided by the radiology departments) was sent to GPs. Feedback contained the number of requests for lumbar spine and knee radiographs made by the whole practice compared with requests made by all GPs in the study.  **Waltz’s Classification:**  *Use evaluative and iterative strategies.*   - Audit and provide feedback.   *Train and educate stakeholders.*   - Distribute educational materials.   **Reminder message group:**  **Description of Implementation Strategy:**  The referral guidelines were sent by post to all GPs.  Educational messages were attached to the reports of every knee or lumbar spine radiograph requested during the 12-month intervention.  **Waltz’s Classification:**  *Train and educate stakeholders.*   - Distribute educational materials.   *Support clinicians*   - Remind clinicians.   **Audit, feedback & Reminder message group:**  **Description of Implementation Strategy:**  This group received a combined intervention of the implementation strategies from the Audit & feedback group and Reminder message group.  **Waltz’s Classification:**  This group received no additional interventions than the Audit & feedback group and Reminder message group combined. | **Description of Implementation Strategy:**  The referral guidelines were sent by post.  **Waltz’s Classification:**  *Train and educate stakeholders.*   - Distribute educational materials. | **Healthcare Professionals:**  *Referral to imaging*   - Number of each radiograph requested per 1000 patients registered with every practice per year. | The effect of educational reminder messages was an absolute change of -1.53 (95% CI: -2.5 to -0.57) for lumbar spine radiographs and of -1.61 (95% CI: -2.6 to -0.62) for knee radiograph requests; these estimates are both relative reductions of about 20%.  The effect of audit and feedback was an absolute change of -0.07 (95% CI: -1.3 to 0.9) for lumbar spine and 0.04 (95% CI: -0.95 to 1.03) knee radiograph requests. Relative reductions were about 1% (knee) and almost no change (lumbar spine).  There was no statistically significant increased effect of receiving both interventions for both types of radiograph |
| Engers et al., 2005 Cluster RCT  The Netherlands | GP’s (67)  Patients with LBP (531)  Primary care | **Description of Implementation Strategy:**  General practitioners received a 2-hour workshop.  Educational material in form of two scientific articles and the guideline, a tool for patient education and reaching agreement on low back pain care was developed and it’s use promoted.  **Waltz’s Classification:**  *Train and educate stakeholders.*   - Develop educational materials. - Make training dynamic. - Distribute educational materials. - Conduct educational meetings.   *Support clinicians*   - Remind clinicians | The control group received no intervention and was labelled as “usual care”. | **Healthcare Professionals:**  *Adherence to implemented intervention.*   - Adequacy of patient education   *Referral to secondary care*   - The number of referrals to a therapists   *Prescription of analgesics*   - Prescription of pain medication | Data showed that the intervention did not promote statistical significance on any outcomes between groups.  The overall number of referrals to a physical, exercise, or manual therapist did not differ for the intervention group compared with the control group.  The advice and explanation provided by the general practitioners, the prescription of paracetamol or NSAIDs, and the prescription of pain medication on a time contingent or a pain contingent basis showed no statistically significant differences between the intervention group and the control group. |
| Evans et al., 2010 Cluster RCT  United Kingdom | Physiotherapists (824); chiropractors (336); and osteopaths (598)  Patients with LBP  Primary care | **Description of Implementation Strategy:**  Printed information package posted to the participants with evidence-based management of acute LBP, based on the latest guidelines.  **Waltz’s Classification:**  *Train and educate stakeholders:*   - Develop educational materials. - Distribute educational materials | No intervention | **Healthcare Professionals:**  *Adherence to implemented intervention.*  Quality indicators reflecting clinical behavior are measured by:   - Activity - Work - Bedrest | Odds of being guideline consistent in the intervention group compared to the control group:   - Activity:  OR 1.286 (95% CI: 0.027 to 1.610) p=0.028 - Work:  OR 1.346 (95% CI: 1.069 to 1.695) p=0.012 - Bed rest: OR 1.308 (95% CI: 0.970 to 1.763) p=0.078 |
| French et al., 2013  Cluster RCT Australia | GPs (112)  Patients with LBP  Primary care (92 practices) | **Description of Implementation Strategy:**  The selected behaviour changes techniques included information provision, persuasive communication, provide information on consequences, provide opportunities for social comparison, barrier identification, model/demonstrate the behaviour, role play, provide instruction, time management and action planning. These techniques were combined and delivered via two facilitated, interactive, educational workshops, each of three hours’ duration. Workshops were a combination of didactic lectures and small group discussions and activities. A DVD was produced and distributed to all GPs in the intervention group**.**  **Waltz’s Classification:**  *Use evaluative and iterative strategies.*   - Assess for readiness and identify barriers and facilitators.   *Provide interactive assistance.*   - Facilitation   *Adapt and tailor to context.*   - Tailor strategies   *Develop stakeholder interrelationships.*   - Conduct local consensus discussions.   *Train and educate stakeholders.*   - Conduct ongoing training. - Make training dynamic. - Distribute educational materials. - Conduct educational meetings | **Description of Implementation Strategy:**  Received a printed copy of the guideline and a written reminder of how to access the electronic version of the guideline.  **Waltz’s Classification:**  *Train and educate stakeholders.*   - Distribute educational materials.   *Support clinicians*   - Remind clinicians. | **Healthcare Professionals:**  *Uptake of knowledge*  Behavioural simulation:   - X-ray referral - Any imaging referral - Advice to stay active. - Advice regarding bed rest   *Referral to imaging*  X-ray and CT rates pr patient seen:   - X-ray referral - CT-Scan referral - X-ray or CT-scan referral | **Behavioural simulation**  X-ray adherence: Adj OR.76 (95% CI: 1.01 to 3.05) p=0.045  Imaging adherence: Adj OR 2.36 (95% CI: 1.48 to 3.79) p=0.000  Activity adherence: Adj. OR 4.49 (95% CI: 1.90 to 10.60) p=0.001  Bed rest adherence: Adj. OR 2.91 (95% CI: 0.30 to 27.83) p=0.354  **X-ray and CT rates pr patient seen.**  X-ray referral: 0.83 (95% CI: 0.61 to 1.12) p=0.211  CT-scan referral: 0.92 (95% CI: 0.66 to 1.27) p=0.598  X-ray or CT-scan referral: 0.87 (95% CI: 0.68 to 1.10) p=0.244 |
| French et al., 2022 Cluster RCT  Australia | Physiotherapists (182); Chiropractors (88)  Patients with LBP (1358)  Primary care (104 practices) | **Description of Implementation Strategy:**  Participated in a full day symposium with both didactic lectures relevant to the guidelines delivered by opinion leaders, along with practical rehearsal sessions. Educational materials were developed and distributed. Clinicians were reminded and support with follow-up phone calls.  **Waltz’s Classification:**  *Use evaluative and iterative strategies*   - Assess for readiness and identify barriers and facilitators   *Adapt and tailor to context*   - Tailor strategies   *Develop stakeholder interrelationships*   - Identify and prepare champions - Inform local opinion leaders - Use an implementation advisor   *Train and educate stakeholders*   - Develop educational materials - Distribute educational materials - Conduct educational outreach visits   *Support clinicians*   - Remind clinicians | **Description of Implementation Strategy:**  A printed copy of the guideline.  A reminder of where to find the guideline.  **Waltz’s Classification:**  *Develop stakeholder interrelationships*   - Inform local opinion leaders   *Train and educate stakeholders*   - Distribute educational materials   *Support clinicians*   - Remind clinicians | **Healthcare Professionals:**  *Referral to imaging*   - Number of patients being referred to x-ray.   **Patients:**  *Function*   - Roland-Morris Disability questionnaire (RMDQ) | There was no statistically significant difference between groups in the odds of patients being referred for X-ray with an adjusted (Adj) OR of 1.40 (95% CI: 0.51 to 3.87, p=0.514) and an Adj risk difference (RD) of 0.01 (95% CI: -0.02 to 0.04).  There was no important clinical difference in low back pain-specific disability between groups with an Adj mean difference of 0.37 (95% CI: -0.48 to 1.21). The estimated ICC for referral for X-ray was 0.15 (95% CI 0.09 to 0.17) and for low back pain-specific disability was 0 (95% CI 0 to 0.07). |
| Goldberg et al., 2001  Cluster RCT  The United States of America | Spinal surgeons  Primary physicians  Patients with LBP  Primary care (10 communities inhabited by 245.710 citizens) | **Description of Implementation Strategy:**  High volume surgeons who were considered opinion leaders were contacted and attended meetings and was expected to share information locally. Continued medical education was utilized in study communities. Academic detailing with posters and pamphlets distributed in the clinics and hospitals.  **Waltz’s Classification:**  *Use evaluative and iterative strategies.*   - Audit and provide feedback.   *Develop stakeholder interrelationships.*   - Inform local opinion leaders. - Conduct local consensus discussions.   *Train and educate stakeholders.*   - Develop educational materials. - Distribute educational materials. - Conduct educational meetings.   *Change infrastructure.*   - Change record systems | The control group received no intervention and was labelled as “usual care”. | **Healthcare Professionals:**  *Adherence to implemented intervention.*   - Quarterly observed rate of surgeries per 100.000 people | The difference between study and control group means were plotted and had a slope of 0.85 in the baseline period, and in the intervention period, a negative slope on -1.24, resulting in a delta slope on -2.09 (p=0.01). |
| Hoeijenbos et al., 2005  Cluster RCT  The Netherlands | Physiotherapist (113)  Patients with LBP (500)  Primary care (68 Practices) | This study was performed using data from Bekkering et al., 2005 (a).  For a description of the implementation strategies and Waltz’s Classification, see Bekkering et al., 2005 (a). | | **Patients:**  *Quality of life*   - Health effects (EQ-5D)   **Cost-effectiveness:**   - Average costs per patient - Average intervention costs per physiotherapist | **EQ-5D Baseline**:  Control: 0.6134 (S.D. = 0.2661)  Intervention: 0.6730 (S.D. = 0.2042)  p=0.006  **6 weeks**  Control: 0.7497 (S.D. = 0.2316)  Intervention: 0.7778 (S.D. = 0.1978)  **12 weeks**  Control: 0.8141 (S.D. = 0.1988)  Intervention: 0.7873 (S.D. = 0.2210)  **Cost-effectiveness:**  **Total average cost per patient at baseline (median)**  Intervention group: € 97(72)  Control group: €89(71)  **Total average cost per patient at 6 weeks (median)**  Intervention group: €125(111)  Control group: €145(141)  **Total average cost per patient at 12 weeks (median)**  Intervention group: €58(20)  Control group: €77(25)  p= 0.051  **Total average cost per patient at 26 weeks (median)**  Intervention group: €33(0)  Control group: €35(0)  p=0.818  **Total average cost per patient at 52 weeks (median)**  Intervention group: €24(0)  Control group: €30(0)  p=0.477 |
| Jensen et al., 2017  Cluster RCT  Denmark. | GP’s  Patients with LBP (475)  Primary care (60 practices) | This study was performed using data from Riis et al., 2016.  For a description of the implementation strategies and Waltz’s Classification, see Riis et al., 2016. | | **Cost-Effectiveness:**  *Health-related quality of life*  From Health-related quality of life (HRQoL) using EuroQoL-5Dimensions-5Level (EQ-5D-5L) the quality and quantity of life lived, was calculated as the area under the curve.  *Intervention costs*  *Health care costs*   - Primary sector - Secondary sector   *Cost–utility analysis*  *Scenario analyses*  quality-adjusted life years (QALYs) | Results showed that costs associated with primary health care were higher, whereas secondary health care costs were lower for the intervention group when compared with the control group.  The total cost of the multifaceted implementation strategies (MuIS) amounted to €34836, equivalent to €65 per patient.  The adjusted cost regression did not show a statistically significant average cost saving per patient of €169 (95% CI: -€661 to €323).  The adjusted QALY decrement regression did not show a statistically significant average loss of 0.0065 QALY (95% CI: -0.036 to 0.023) per patient over the 12 months of follow-up. |
| Leonhardt et al., 2008  RCT  Germany | GP’s (126)  Patients with LBP (1378)  Primary care (118 practices) | This study was performed using data from Becker et al., 2008.  For a description of the implementation strategies and Waltz’s Classification, see Becker et al., 2008. | | **Patients:**  *Physical activity*   - Freiburger Questionnaire on physical activity (FQPA) | **Total physical activity score compared to controls**:  At 6 months  Group A: MD 2.95 (95% CI: -1.64 to 7.54) p=0.21  Group B: MD 2.77 (95% CI: -1.81 to 7.34) p=0.23  At 12 months  Group A: MD 3.58 (95% CI: -1.42 to8.59) p=0.16  Group B: MD 2.57 (95% CI: -2.43 to7.56) p=0.31 |
| Maas et al., 2015 Cluster RCT  The Netherlands | Physiotherapists (149)  Patients with LBP  Communities of practice | **Peer assessment (PA) group:**  **Description of Implementation Strategy:**  Received a link to the KNGF guidelines.  Received by e-mail a program guide tailored to the intervention providing detailed information about learning objectives, learning content, training schedule, didactic format, and procedure.  The program consisted of four 3-hour sessions: In three of the sessions the participants worked on written cases. Session 3 consisted of a review of patient records and PA.  **Waltz’s Classification:**  *Provide interactive assistance.*   - Provide clinical supervision.   *Train and educate stakeholders.*   - Conduct ongoing training. - Make training dynamic. - Distribute educational materials. - Conduct educational meetings | **Case discussion (CD) group:**  **Description of Implementation Strategy:**  This group received the same material but focused on discussion of the material instead of peer assessment. Session 3 consisted of a review of patient records and CD.  **Waltz’s Classification:**  The CD group received the same interventions as det PA group except “Provide clinical supervision” and instead received:  *Develop stakeholder interrelationships.*   - Conduct local consensus discussions | **Healthcare Professionals:**  *Uptake of knowledge*  **Guideline adherence:**   - Online test/questionnaire based on 4 vignettes.   **Reflective practice:**   - Self-Reflection and Insight Scale (SRIS)   **Awareness of performance:**   - Awareness was conceived of as the association between perceived improvement and assessed improvement. | **Guideline adherence:**  Difference between the PA and CD groups change: Estimated difference of 22.52 points (95% CI: 2.38 to 42.66, p=0.031) in favour of the PA group.  **Reflective practice:**  Difference between the PA and CD groups change: Estimated difference of -0.06 points (95% CI: -2.79 to 2.65, p=0.96) in favour of the PA group  **Awareness of performance:**  Difference between the PA and CD groups change: Estimated difference 14.73 (95% CI: 2.78 to 26.68, p=0.01)) in favour of the PA group |
| Mortimer et al., 2013  Cluster RCT  Australia | GP’s (112)  Patients with LBP  Primary care (92 practices) | This study was performed using data from French et al., 2013.  For a description of the implementation strategies and Waltz’s Classification, see French et al., 2013. | | **Cost-Effectiveness:**  *Cost of development and delivery*  *Cost effectiveness of delivery*  *Cost effectiveness of development and delivery* | **Cost-effectiveness:**  Adjusted incidence rate ratios suggest that total cost per GP was 0.92 times lower in the intervention group than control group GPs but not statistically significant (p = 0.578).  Incremental effects showed that exposure to the intervention reduced total cost per GP by $375.55 but was not statistically significant (95% CI: -$1815.63, $1064.53; p = 0.605). |
| Moseng et al., 2019 Stepped wedge cluster RCT.  Norway | GP’s (40)  Physiotherapists (37)  Patients with hip and knee OA (393)  Primary care | **Description of Implementation Strategy:**  One day seminar who included:   - General update on OA epidemiology, clinical features and treatment recommendations. - Education in how to deliver a standardized patient-education programme. - Education in delivering individualized exercise intervention and performance tests. - Education about nutrition and weight management.   **Waltz’s Classification:**  *Train and educate stakeholders.*   - Distribute educational materials. - Conduct educational meetings. - Conduct educational outreach visits | No intervention because of a Stepped wedge design | **Patients:**  *Patient adherence*   - Uptake of recommended core treatment modalities in OA | **Findings:**  Patient-reported data showed a statistically significant higher uptake for all the core treatment components for the intervention group compared to the control group (p<0.05).  Seven out of eight items had a guideline consistency of ≥ 90%.  Ratio for patients in the intervention period participated in patient education, compared to the control period:   - Adj OR: 82.2; 95% CI: 24.6 to 274.7 p<0.001   Ratio for patients in the intervention period performed resistance exercise, compared to the control period:   - Adj OR: 5.0; 95% CI: 2.1 to 12.3 p<0.001   Ratio for patients in the intervention period performed cardiorespiratory exercise, compared to the control period:   - Adj OR: 5.7; 95% CI: 2.5 to 13.1 P<0.001 |
| Murray et al., 2015  Cluster RCT  Ireland | Physiotherapists (24**)**  Patients with LBP (24)  Secondary care | **Description of Implementation Strategy:**  Physiotherapists attended a 1-hour education session. This session reviewed current best evidence-based care for CLBP management.  Physiotherapists in the intervention group also participated in two sessions 4-h sessions of communication skills training.  The two sessions consisted of strategies for implementing SDT, video recordings of vignettes, active roleplay and group discussions including facilitators and barriers for implementation.  Individualized e-mails were sent to the physiotherapists to discuss progress toward attainment of implementation goals and assistance in resolving any problems.  **Waltz’s Classification:**  *Develop stakeholder interrelationships.*   - Conduct local consensus discussions.   *Train and educate stakeholders.*   - Make training dynamic. - Distribute educational materials. - Conduct educational meetings | **Description of Implementation Strategy:**  The control got the same 1-hour education session as the intervention group.  **Waltz’s Classification:**  *Train and educate stakeholders.*   - Conduct educational meetings | **Healthcare Professionals:**  *Adherence to implemented intervention.*  Physiotherapists’ needs-supportive communication: Raters each listened to 12 recordings from initial treatment sessions with a patient and used the Health Care Climate Questionnaire (HCCQ) to assess physiotherapists’ needs-supportive communication. | Between group difference in favour of intervention group (HCCQ): d=2.27 (95% CI: 1.18 to 3.21, p<0.001) |
| O'Connor et al., 2022  Cluster RCT  Australia | GP’s (3819)  Patients with musculoskeletal conditions  Primary care | **Description of Implementation Strategy:**  General practitioners received individualized written audit and feedback. Four different methods of using audit and feedback were applied, and compared both with each other, and with the control group. The audit and feedback were either in an enhanced visual display, or a standard one, and either once or twice in the two different audit and feedback group, thus giving 4 groups.  **Waltz’s Classification:**  *Use evaluative and iterative strategies.*   - Audit and provide feedback.   *Train and educate stakeholders.*   - Distribute educational materials | No intervention. | **Healthcare Professionals:**  *Referral to imaging*  Overall rate of imaging requests per 1000 patient consultations   - In intervention groups compared to control - Difference between intervention groups | Rate of imaging referrals in the intervention groups compared to the control: Adj MD −2.66 (95% CI: −3.24 to −2.07, p<0.001) |
| Peter et al., 2013 RCT  The Netherlands. | Physiotherapists (248)  Patients with hip and knee OA  Private practice | **Description of Implementation Strategy:**  The interactive workshop, which consisted of two educational courses, was led by an expert physiotherapist and 3-4 physiotherapist teachers who received 1.5h instructions about the workshop. The workshop started with a short summary of guideline recommendations. During the workshop physiotherapists examined real patients in small groups. During this process the expert was available to give additional feedback. The workshop ended with a discussion about a fictional case.  **Waltz’s Classification:**  *Provide interactive assistance.*   - Provide clinical supervision.   *Develop stakeholder interrelationships.*   - Identify and prepare champions. - Conduct local consensus discussions.   *Train and educate stakeholders.*   - Make training dynamic. - Conduct educational meetings | **Description of Implementation Strategy:**  The conventional education was a presentation based on the guideline development process and the recommendation in the guideline, along with two different cases.  **Waltz’s Classification:**  *Train and educate stakeholders.*   - Conduct educational meetings | **Healthcare Professionals:**  *Adherence to implemented intervention.*  Adherence to implemented OA guideline (Quality indicators for physical therapy in hip and knee osteoarthritis [QIP-HKOA])  *Uptake of knowledge*  Measured knowledge uptake on a self-made questionnaire. | **Adherence to implemented intervention:**  Change in intervention group (QIP-HKOA):   - MD 2.0 (95% CI: 1.1 to 2.9) from baseline to immediately after. - MD 1.4 (95% CI: 0.6 to 2.2) from immediately after to three months after.   Change in control:   - MD 0.7 (95% CI: -0.3 to 1.7) from baseline to immediately after. - MD 0.9 (95% CI: 0.1 to 2.1) from immediately after to three months after.   Difference between groups: p=0.024  **Uptake of knowledge**  Change in intervention group.   - MD 5.0 (95% CI: 3.4 to 6.6) from baseline to immediately after. - MD -0.7 (95% CI: -2.4 to 0.3) from immediately after to three months after.   Change in control:   - MD 4.4 (95% CI: 2.6 to 6.1) from baseline to immediately after. - MD -2.0 (95% CI: -4.1 to 0.3) from immediately after to three months after.   Difference between groups: p=0.278 |
| Peter et al., 2015 RCT  The Netherlands | Physiotherapists (319)  Patients with hip and knee OA  Primary care | **Description of Implementation Strategy:** The intervention workshop comprised of two educational courses and the interactive, educational course was guided by an expert physiotherapist, in cooperation with 3-4 patients and 3-4 physiotherapy teachers, who were instructed concerning their role.  The workshop started with a short summary of guideline recommendations. During the workshop physiotherapist examined real patients in small groups. During this process, the expert was available to give additional feedback. The workshop ended with a discussion about a fictional case.  **Waltz’s Classification:**  *Provide interactive assistance.*   - Provide clinical supervision.   *Develop stakeholder interrelationships.*   - Identify and prepare champions. - Conduct local consensus discussions.   *Train and educate stakeholders.*   - Make training dynamic. - Conduct educational meetings | No intervention.  The control group received the same educational course 4 months after the first course in every region. | **Healthcare Professionals:**  *Uptake of knowledge*  Self-reported adherence and knowledge of the recommendations in the guideline (Self-developed questionnaire) | **Adherence questionnaire (total score range 0–24):**  Between group difference:  Difference at T1 MD 1.4 (95% CI: 0.7 to 2.0, p<0.001)  Difference at T2 MD 0.9 (95% CI: 0.2 to1.7, p=0.004)  **Knowledge questionnaire (total score range 0–76):**  Between group difference:  Difference at T1 MD 6.8 (95% CI: 4.5 to 9.1, p<0.001)  Difference at T2 MD 3.9 (95% CI: 1.7 to 6.2, p=0.004) |
| Rebbeck et al., 2006  Cluster RCT Australia | Physiotherapists (27)  Patients with whiplash (103)  Primary care | **Description of Implementation Strategy:**  Physiotherapists attended an 8-hour interactive workshop outlining the content of the guidelines and included interactive- and practical sessions. Local opinion leaders delivered some of the content. Furthermore, had follow-up educational outreach visits for 2-hours 6 months later.  **Waltz’s Classification:**  *Develop stakeholder interrelationships.*   - Inform local opinion leaders.   *Train and educate stakeholders.*   - Conduct ongoing training. - Develop educational materials. - Make training dynamic. - Distribute educational materials. - Conduct educational meetings. - Conduct educational outreach visits | **Description of Implementation Strategy:**  Control group received a mail with the guidelines.  **Waltz’s Classification:**  *Train and educate stakeholders.*   - Distribute educational materials. | **Healthcare Professionals:**  *Adherence to implemented intervention.*   - Clinical practice (Percentage of physiotherapists prescribing guideline recommendations)   *Uptake of knowledge*   - Custom-made questionnaire (range 0-28)   *HCP satisfaction*   - Physiotherapist satisfaction (7-point Likert scale)   **Patients:**  *Function*   - Functional Rating Index   *Patient Satisfaction*   - Patient satisfaction (5-point Likert scale) - Global Perceived Effect | **Health care professionals**  **Uptake of knowledge:**   - Knowledge uptake: 5.5 points (95% CI: 2.5 to 8.4, p = 0.001) compared to the control   Self-rated guideline understanding: 1.5 points (95% CI: 0.7 to 2.3, p = 0.001) compared to the control  **Adherence to implemented intervention:**  Specific guideline consistent advice “advise to act as usual”:   - Intervention group: 7% to 67% increase. - Control group: 8% before, and 18% increase.   **Prescribing according to guideline recommendation after trial:**  **Reassure patient** p = 0.05 **Advise to act as usual** p= 0.04 **Prescribe function** p= 0.22 **Prescribe exercise** p= 1.00 **Prescribe medication** p= 0.10  **Prescribing 2 of 5 guideline recommendation:**   - Intention: 44% - Actual: 32%   **Patients (difference between groups)**  **Functional rating index:**  MD –0.6 (95% CI: –7.8 to 6.6) p = 0.87  **Core outcome measure**  MD 0.3 (95% CI: –2.4 to 3.0) p = 0.85  **Global perceived effect.**  MD 0.1 (95% CI: –1.7 to 1.8) p = 0.95  **Satisfaction:**  GP Care p=0.69, Physiotherapy care p=0.87, Consumer version of guidelines p =0.93 |
| Riis et al., 2016  Cluster RCT  Denmark | GP’s (60)  Patients with LBP (1101)  Primary care | **Description of Implementation Strategy:**  Usual implementation activity, which were a regional information meeting about the guidelines. In addition, the MuIS group had outreach visits by a primary care physiotherapist, who were trained to convey the content of the LBP guidelines, and practices were offered follow-up with the outreach visitor.  **Waltz’s Classification:**  *Develop stakeholder interrelationships.*   - Conduct local consensus discussions.   *Train and educate stakeholders.*   - Conduct ongoing training. - Provide ongoing consultation. - Distribute educational materials. - Conduct educational meetings. - Conduct educational outreach visits.   *Support clinicians*   - Remind clinicians.   *Engage consumers.*   - Use mass media.   *Change infrastructure.*   - Change record systems | **Description of Implementation Strategy:**  Usual implementation activity, which were a regional information meeting about the guidelines.  **Waltz’s Classification:**  *Train and educate stakeholders.*   - Conduct ongoing training. - Distribute educational materials. - Conduct educational meetings.   *Engage consumers.*   - Use mass media. - *Change infrastructure* systems | **Healthcare Professionals:**  *Referral to secondary care*   - Number of patients referred to secondary care. | **Referrals to secondary care:**   - 27 (5%) in the intervention group - 59 (10.5%) in the passive group, - Adj. OR 0.52 (CI: 95% 0.30 to 0.90, p = 0.020) |
| Sanders et al., 2017  Cluster RCT  The Netherlands | GP’s (42)  Patients with LBP (226)  Primary care | **Description of Implementation Strategy:**  GPs in the intervention group received two training sessions of two and a half hours in duration and were held in small groups. The training sessions included, group discussion, theory, role-playing and reflections on personal behaviour were alternated.  A decision aid for non-chronic, non-specific low back pain was developed based on the internationally accepted IPADS guidelines. To stimulate their use of SDM skills during the actual consultations, we provided the GPs with a desktop tool containing group-formulated open-ended questions applicable to the consecutive SDM process elements. In addition to receiving training sessions, the GPs in the intervention group received personalized feedback on two videotaped consultations.  **Waltz’s Classification:**  *Provide interactive assistance.*   - Provide clinical supervision.   *Develop stakeholder interrelationships.*   - Conduct local consensus discussions.   *Train and educate stakeholders.*   - Develop educational materials. - Make training dynamic. - Distribute educational materials. - Conduct educational meetings | The control group received no intervention and was labelled as “usual care”. | **Healthcare Professionals:**  *Adherence to implemented intervention.*   - Level of SDM (the OPTION scale) - Level of positive reinforcement of the chosen therapy (Positive reinforcement of the chosen therapy) | **Level of SDM (OPTION Scale):**   - MD 14.86 (p<0.05)   **Level of positive reinforcement:**   - MD 0.77 (p<0.05) |
| Schectman et al., 2003  Cluster RCT  The United States of America | Clinicians (85)  Patients with LBP (2020)  Primary care | **Description of Implementation Strategy:**  A guideline was developed, along with educational materials and distributed. Received a 90-minute educational outreach with a didactic outlining of the guideline, and a series of interactive educational vignettes, and were delivered by local opinion leaders. Each clinician received audit and feedback and a follow-up phone call and visit. Received writing reminders twice in the additional months of the study year. Educational materials were both distributed to clinicians and patients.  **Waltz’s Classification:**  *Use evaluative and iterative strategies.*   - Audit and provide feedback.   *Develop stakeholder interrelationships:*   - Inform local opinion leaders.   *Train and educate stakeholders.*   - Develop educational materials. - Distribute educational materials. - Conduct educational outreach visits.   *Support clinicians*   - Remind clinicians | No intervention | **Healthcare Professionals:**  *Referral to imaging*  The utilization of imaging investigations:   - X-ray - CT or MR   *Referral to secondary care*   - Physical therapy referrals - Subspecialty referral | **Overall**  Guideline-consistent behavior increased by 5.4% in the intervention group. Declines 2.7% in the control group (p = 0.046).  Overall decline in raw utilization of services of 8.5% in the intervention group versus 0.6% in the control group (p = 0.042).  **X-ray referrals:**  Intervention group:   - Baseline: 31% (14.5 not guideline consistent) - Intervention year: 19% (8.1% not guideline consistent)   Control group:   - Baseline: 21% (8.2% not guideline consistent)   Intervention year: 18% (8.6% not guideline consistent)  **CT or MRI referrals:**  Intervention group:   - Baseline: 7.6% (5.7% not guideline consistent) - Intervention year: 5.6% (3.5% not guideline consistent)   Control group:   - Baseline: 5.6% (3.5% not guideline consistent) - Intervention year: 7.1% (5.4% not guideline consistent)   **PT referrals:**  Intervention group:   - Baseline: 12% (10% not guideline consistent) - Intervention year: 10% (9.2% not guideline consistent)   Control group:   - Baseline: 13% (10.9% not guideline consistent) - Intervention year: 13% (12% not guideline consistent)   **Specialty referrals:**  Intervention group:   - Baseline: 12% (9.5% not guideline consistent) - Intervention year: 8.6% (7.1% not guideline consistent)   Control group:   - Baseline: 5.9% (4.0% not guideline consistent) - Intervention year: 7.1% (5.6% not guideline consistent) |
| Scheel et al. 2002  Cluster RCT  Norway | GP’s  Patients with LBP (6179)  Primary care (65 municipalities) | **Passive strategy group:**  **Description of Implementation Strategy:**  Passive implementation strategies, which were both low in cost and quick to implement: Targeted information to GP’s, a checkbox in the form for reporting sick leave, which served as a reminder, a desktop summary of the clinical guidelines.  **Waltz’s Classification:**  *Use evaluative and iterative strategies.*   - Assess for readiness and identify barriers and facilitators.   *Train and educate stakeholders.*   - Develop educational materials. - Distribute educational materials.   *Support clinicians*   - Remind clinicians     **Proactive strategy group:**  **Description of Implementation Strategy:**  In addition to the passive group, they also received:  Resource person to provide proactive support to GP’s and actively following patients on sick leave for less than 16 days, and a workshop on LBP and ASL.  **Waltz’s Classification:**  In addition to the passive strategy group, the proactive strategy group received:  *Provide interactive assistance.*   - Facilitation   *Develop stakeholder interrelationships.*   - Inform local opinion leaders.   *Train and educate stakeholders.*   - Conduct ongoing training. - Provide ongoing consultation. - Conduct educational meetings | No intervention. | **Healthcare Professionals:**  *Adherence to implemented intervention*  Measuring the total utilization of active sick leave.   - The total number of days on sick leave - The number of days on ordinary sick leave - The number of days on ordinary sick leave before starting ASL | **Net increase usage of Active Sick Leave:**   - Proactive group (17.7%) - Passive group (10.8%) - Control group (12.4%) - Cluster-adjusted χ2 5.67, df = 2, (p= 0.018) |
| Schröder et al., 2022  Stepped cluster RCT.  Sweden | Physiotherapists (98)  Patients with LBP (500)  Primary care | **Description of Implementation Strategy:**  The BetterBack Model of Care (MoC) support team designed the mandatory 2-day workshop (13.5-hour) for the physiotherapists. The workshop started with a presentation of the local adaptation process together with the 11 guideline recommendations and thereafter presentation and practical use of clinical tools to support CPG adherent health-care delivery. The workshop also involved patient cases, where physiotherapists worked in small groups.  **Waltz’s Classification:**  *Provide interactive assistance.*   - Facilitation   *Train and educate stakeholders.*   - Conduct ongoing training. - Make training dynamic. - Distribute educational materials. - Conduct educational meetings | The control group received no intervention and was delivering routine care. | **Healthcare Professionals:**  *Referral to secondary care*   - Referrals to specialist consultation   *Referral to imaging*   - Referrals to medical imaging | **Between group comparison:**   - Received referral to specialist consultation: OR 06 (95% CI: 0.3 to 1.4) p=0.257: ICC = 0 - Received medical imaging: OR 0.5 (95% CI: 0.3 to 0.8); p = 0.011: ICC < 0.001 |
| Simula et al., 2021  Cluster RCT  Finland | Physiotherapists, nurses and physicians  Patients with LBP (415)  Primary care (8 Health centers) | **Description of Implementation Strategy:**  A booklet was developed and distributed, along with a 30 min session on its use.  **Waltz’s Classification:**  *Train and educate stakeholders.*   - Develop educational materials. - Distribute educational materials. - Conduct educational meetings | No intervention. | **Healthcare Professionals:**  *Referral to imaging*  Total number of imaging investigations:   - The proportion of patients presenting with LBP who underwent imaging examinations due to LBP.   **Patients:**  *Function*   - Change in Patient-Reported Outcomes Measurement Information System, 20-item physical functioning short form (PROMIS PF-20) | **Referrals to imaging at 12 months:**   - Adj. OR (95% CI: 0.52 (0.32 to 0.84 p=0.007)   **Referrals to radiographs at 12 months:**   - Adj. OR 0.58 (95% CI: 0.37 to 0.90) p=0.016   **Referrals to MRI at 12 months:**   - Adj. OR 0.64 (95% CI: 0.49 to 0.84) p=0.001   **Referrals to MRI + CT at 12 months:**   - Adj. OR 0.64 (95% CI: 0.45 to 0.84) p=0.014   **PROMIS T-Score difference at 12 months:**   - Adj. MD 0.09 (95% CI: -2.3 to 2.3) p=0.935 |
| Stevenson et al., 2004  Cluster RCT England | Physiotherapists (30)  Patients with LBP  Community Trust | **Description of Implementation Strategy:**  The intervention group received an interactive evidence-based educational program, including the use of opinion leaders, teaching, discussion, reflective thinking, active experimentation, and peer group teaching. Participants received a two and half-hour session on critical appraisal and literature searching skills and a two and a half-hour session on the latest management for patients with acute and chronic low back pain.  **Waltz’s Classification:**  *Develop stakeholder interrelationships.*   - Inform local opinion leaders. - Conduct local consensus discussions.   *Train and educate stakeholders.*   - Conduct ongoing training. - Make training dynamic. - Conduct educational meetings | **Description of Implementation Strategy:**  The control group received a standard in-service training package on clinical management of knee dysfunction and pathology.  The training consisted of two sessions, each lasting two and a half hours and involved a discussion of the subjective and objective examination of the knee complex.  **Waltz’s Classification:**  *Develop stakeholder interrelationships.*   - Conduct local consensus discussions.   *Train and educate stakeholders.*   - Conduct ongoing training. - Make training dynamic. - Conduct educational meetings | **Healthcare Professionals:**  *Uptake of knowledge*  Change in physiotherapists’ attitudes towards evidence-based practice, concerning following domains:(Self-administered questionnaire)   - Clinical practice should be based on the best available evidence. - We should change our practice if good quality evidence suggests we should. - I would find it difficult to change what I already do in clinical practice. - I have support from management to undertake EBP. - I would lack confidence in undertaking a literature search. - I would feel confident in undertaking a critical appraisal. - Journal articles are easy to read. | **Therapist has support to undertake EPB:**  Intervention group:   - Baseline 47% - 6 months: 81%   Control group:   - Baseline: 38% - 6 months: 45%   **Confidence in critical appraisal:**  Intervention group:   - Baseline 24% - 6 months: 62%   Control group:   - Baseline: 15% - 6 months: 50% |
| Stevenson et al., 2006  Cluster RCT England | Physiotherapists (30)  Patients with LBP (306)  Primary care | This study was performed using data from Stevenson et al., 2004.  For a description of the implementation strategies and Waltz’s Classification, see Stevenson et al., 2004. | | **Healthcare Professionals**  *Adherence to implemented intervention.*  Guideline consistent practice advice, measured by discharge summaries, as measured on five different items of guideline consistent advice to patients.  *Uptake of knowledge*  Alteration of attitudes and beliefs about pain. | **Guideline consistent advice:**   - Advice about work situation: OR 1.1 (95% CI: 0.4 to 2.6) - Advice on return to normal activities: OR 0.9 (95% CI: 0.3 to 2.8) - Advice to increase activity level: OR 1.2 (95% CI: 0.3 to 4.4) - Encourage early return to work: OR 2.0 (95% CI: 0.1 to 34.5) - Encourage to undertake activities themselves: OR 0.2 (95% CI: 0.0 to 0.7)   **Attitudes and beliefs:**   - Change attitudes/beliefs about pain: OR 2.3 (95% CI: 0.6 to 9.3). |
| Suman et al., 2018 Cluster RCT  The Netherlands | GP’s (53)  Patients with LBP (5130)  Primary care (25 practices) | **Description of Implementation Strategy:**  Multidisciplinary continuing medical education in communications interprofessionally and with patients were main themes as means to reduces referral, done through distribution of educational materials, and via an interactive website.  **Waltz’s Classification:**  *Train and educate stakeholders*   - Conduct ongoing training - Distribute educational materials - Conduct educational meetings   *Engage consumers*   - Use mass media | The control group received no intervention and was delivering usual care. | **Healthcare Professionals:**  Guideline adherence was assessed using performance indicators:  *Referral to imaging*   - Referral for diagnostic imaging   *Referral to secondary care.*   - Referral to consultation with medical specialists (neurology, orthopedics or other specialty) - Referral for psychosocial care as indicator for multidisciplinary collaboration - Referral to and/or contact with occupational physician as indicator for multidisciplinary collaboration | No significant difference between groups over time on the number of total referrals to imaging and medical specialist care (p>0.05).  **Referrals to medical imaging:**   - Baseline: 200 (14%) - Follow-up: 138 (11%)   Control group:   - Baseline: 145 (12%)   **Referrals to medical specialists:**  Intervention group:   - Baseline: 171 (12%) - Follow-up: 100 (8%)   Control group:   - Baseline: 109 (9%) - Follow-up: 99 (8%) - Follow-up: 137 (11%) |
| Van Dulmen et al., 2014  Cluster RCT  The Netherlands | Physiotherapists (90)  Patients with LBP  Communities of practice | **Problem-Based Peer Assessment (PA):**  **Description of Implementation Strategy:**  The educational approaches consisted of a series of four 2-hour meetings during a 6-month period.  In the Peer Assessment group, clinical performance was directly observed and evaluated by peers in a simulated setting. Participants performed in 3 roles: physical therapist, assessor, and patient.  Participants received a peer-assessment manual in advance.  The expert assessor provided additional feedback only if necessary and when all peers had given their feedback.  **Waltz’s Classification:**  *Provide interactive assistance.*   - Provide clinical supervision.   *Develop stakeholder interrelationships.*   - Conduct local consensus discussions.   *Train and educate stakeholders*   - Conduct ongoing training - Make training dynamic - Distribute educational materials - Conduct educational meetings | **Case-Based Discussion: (CD)**  **Description of Implementation Strategy:**  The educational approaches consisted of a series of four 2-hour meetings during a 6-month period.  In the Case-Base Discussion group, the participants received a program manual and the sessions involved case-based discussions.  **Waltz’s Classification:**  *Develop stakeholder interrelationships.*   - Conduct local consensus discussions.   *Train and educate stakeholders.*   - Conduct ongoing training. - Make training dynamic. - Distribute educational materials. - Conduct educational meetings | **Healthcare Professionals:**  *Uptake of knowledge*   - Therapist knowledge and clinical reasoning (Self-made questionnaires 0-100)   12 quality indicators (Mean scores):   1. “Red flags” assessed correctly. 2. Assessment of the patient’s complaints 3. Correct choice of the patient profile 4. Contacting the physician in case of red flags 5. Choice of examination objectives related to the patient profile. 6. Choice of treatment objectives related to the patient profile. 7. Choice of treatment strategies related to the patient profile. 8. Number of intervention sessions 9. Adequate information is provided. 10. Health outcome questionnaires have been applied. 11. Written report to physician 12. Aftercare has been arranged | **Scores are in favor of PA.**  **Total scores on vignettes:**  8.7% (95% CI: 3.9 to 13.4, p=.001)  **Quality indicators:**  **“Red flags” assessed correctly:**  6.7 (95% CI: 0.3 to 13.1) p=0.04  **Correct choice of the patient profile:**  13.1 (95% CI: 2.2 to 24.0) p=0.02  **Contacting the physician in case of red flags:**  6.8 (95% CI: 1.1 to 12.4) p=0.02  **Choice of treatment strategies related to the patient profile:**  12.4 (95% CI: 1.5 to 23.3) p=0.03 |
| **Abbreviations:** GP: General practitioners; LBP: Low back pain; NSAID: Non-steroidal anti-inflammatory drug; QALY: Quality adjusted life years; MRI: Magnetic resonance imaging; CT: Computerized tomography; EBP: Evidence based practice.  EE: Effect estimate; Adj: adjusted; OR: Odds Ratio; RR: Risk ratio; MD: Mean Difference; RD: Risk difference; β6: Step change; β7: Change in trend; CI: Confidence interval; df: Degrees of freedom; | | | | | |
