## Supplementary material for "Effectiveness of Implementation Interventions in Musculoskeletal Healthcare: A Systematic Review": Synthesis of results

### Additional file 5 - Synthesis of Results - Outcomes for each Implementation Intervention

| ***Table A5.1***  *Synthesis of Results - Outcomes for each Implementation Intervention* | | | | | | | |
| --- | --- | --- | --- | --- | --- | --- | --- |
| **Waltz’s Classification Clusters (no. studies)** | **Implementation Intervention (no. studies)** | **Sample size (n)** | **Outcome domain** | **Positive statistically significant effect in favour of intervention group** | **No positive statistically significant effect in favour of intervention group** | **Mixed findings of statistically significant effect** | **Unknown statistically significant effect** |
| **Use evaluative and iterative strategies (13)** | Assess for readiness and identify barriers and facilitators (9) | Healthcare Professionals (777)  Patients (15,593) | *Adherence to implemented intervention* | Scheel et al., 2002 | Dey et al., 2004 |  |  |
|  |  |  | *Uptake of knowledge* |  |  | French et al., 2013 |  |
|  |  |  | *Referral to imaging* |  | Coombs et al., 2021; Dey et al., 2004; French et al., 2013; French et al., 2022 |  |  |
|  |  |  | *Prescription of analgesics* |  | Dey et al., 2004 |  |  |
|  |  |  | *Referral to secondary care* |  | Dey et al., 2004 |  |  |
|  |  |  | *Function* | Becker et al., 2008 | French et al., 2022 |  |  |
|  |  |  | *Physical activity* |  | Leonhardt et al., 2008 |  |  |
|  |  |  | *Cost-effectiveness* | Becker et al., 2012 | Mortimer et al., 2013 |  |  |
|  | Audit and provide feedback (5) | Healthcare Professionals (4,335)  Patients (8,204) | *Adherence to implemented intervention* | Goldberg et al., 2001 |  |  |  |
|  |  |  | *Referral to imaging* | O'Connor et al., 2022; Schectman et al., 2003 | Coombs et al., 2021; Eccles et al., 2001 |  |  |
|  |  |  | *Referral to secondary care* | Schectman et al., 2003 |  |  |  |
| **Provide interactive assistance (18)** | Facilitation (5) | Healthcare Professionals (210)  Patients (8,866) | *Adherence to implemented intervention* | Scheel et al., 2002 | Dey et al., 2004 |  |  |
|  |  |  | *Uptake of knowledge* |  |  | French et al., 2013 |  |
|  |  |  | *Referral to imaging* | Schröder et al., 2022 | Dey et al., 2004; French et al., 2013 |  |  |
|  |  |  | *Referral to secondary care* |  | Dey et al., 2004; Schröder et al., 2022 |  |  |
|  |  |  | *Prescription of analgesics* |  | Dey et al., 2004 |  |  |
|  |  |  | *Cost-effectiveness* |  | Mortimer et al., 2013 |  |  |
|  | Provide clinical supervision (13) | Healthcare Professionals (1,140)  Patients (3,461) | *Adherence to implemented intervention* | Bekkering et al., 2005 (b) Peter et al., 2013; Sanders et al., 2017 |  |  |  |
|  |  |  | *Uptake of knowledge* | Peter et al., 2015; Van Dulmen et al., 2014 | Chipchase et al., 2016; Peter et al., 2013 | Maas et al., 2015 |  |
|  |  |  | *Function* | Becker et al., 2008; Cleland et al., 2009 | Bekkering et al., 2005 (b); Chipchase et al., 2016 |  |  |
|  |  |  | *Pain* |  | Bekkering et al., 2005 (b), Cleland et al., 2009 |  |  |
|  |  |  | *Workability* |  |  |  | Bekkering et al., 2005 (b) |
|  |  |  | *Physical activity* |  | Leonhardt et al., 2008 |  |  |
|  |  |  | *Quality of life* |  | Hoeijenbos et al., 2005 |  |  |
|  |  |  | *Cost-effectiveness* | Becker et al., 2012 | Hoeijenbos et al., 2005 |  |  |
| **Adapt and tailor to context (5)** | Tailor strategies (5) | Healthcare Professionals (651)  Patients (8,036) | *Adherence to implemented intervention* |  | Dey et al., 2004 |  |  |
|  |  |  | *Uptake of knowledge* |  |  | French et al., 2013 |  |
|  |  |  | *Referral to imaging* |  | Coombs et al., 2021; Dey et al., 2004; French et al., 2013; French et al., 2022 |  |  |
|  |  |  | *Referral to secondary care* |  | Dey et al., 2004 |  |  |
|  |  |  | *Prescription of analgesics* |  | Dey et al., 2004 |  |  |
|  |  |  | *Function* |  | French et al., 2022 |  |  |
|  |  |  | *Cost-effectiveness* |  | Mortimer et al., 2013 |  |  |
| **Develop stakeholder interrelationships (26)** | Identify and prepare champions (3) | Healthcare Professionals (837)  Patients (1358) | *Adherence to implemented intervention* | Peter et al., 2013 |  |  |  |
|  |  |  | *Uptake of knowledge* | Peter et al., 2015 | Peter et al., 2013 |  |  |
|  |  |  | *Referral to imaging* |  | French et al., 2022 |  |  |
|  |  |  | *Function* |  | French et al., 2022 |  |  |
|  | Inform local opinion leaders (7) | Healthcare Professionals (442)  Patients (9966) | *Adherence to implemented intervention* | Goldberg et al., 2001; Rebbeck et al., 2006; Scheel et al., 2002 | Stevenson et al., 2006 |  |  |
|  |  |  | *Uptake of knowledge* | Rebbeck et al., 2006 | Stevenson et al., 2006 |  | Stevenson et al., 2004 |
|  |  |  | *Referral to imaging* | Schectman et al., 2003 | French et al., 2022 |  |  |
|  |  |  | *Referral to secondary care* | Schectman et al., 2003 |  |  |  |
|  |  |  | *HCP satisfaction* |  | Rebbeck et al., 2006 |  |  |
|  |  |  | *Function* |  | French et al., 2022; Rebbeck et al., 2006 |  |  |
|  |  |  | *Patient satisfaction* |  | Rebbeck et al., 2006 |  |  |
|  | Conduct local consensus discussions (22) | Healthcare Professionals (1,496)  Patients (5,880) | *Adherence to implemented intervention* | Bekkering et al., 2005 (a); Goldberg et al., 2001; Murray et al., 2015; Peter et al., 2013; Sanders et al., 2017 | Dey et al., 2004; Stevenson et al., 2006 |  |  |
|  |  |  | *Uptake of knowledge* | Peter et al., 2015; van Dulmen et al., 2014 | Bussières et al., 2010; Chipchase et al., 2016; Peter et al., 2013; Stevenson et al., 2006 | French et al., 2013 | Stevenson et al., 2004 |
|  |  |  | *Referral to imaging* |  | Dey et al., 2004; French et al., 2013 |  |  |
|  |  |  | *Referral to secondary care* | Riis et al., 2016 | Dey et al., 2004 |  |  |
|  |  |  | *Prescription of analgesics* |  | Dey et al., 2004 |  |  |
|  |  |  | *Function* | Becker et al., 2008 | Chipchase et al., 2016; Bekkering et al., 2005 (b); Becker et al., 2008 |  |  |
|  |  |  | *Pain* |  | Bekkering et al., 2005 (b) |  |  |
|  |  |  | *Workability* |  |  |  | Bekkering et al., 2005 (b) |
|  |  |  | *Physical activity* |  | Leonhardt et al., 2008 |  |  |
|  |  |  | *Quality of life* |  | Hoeijenbos et al., 2005 |  |  |
|  |  |  | *Cost-effectiveness* | Becker et al., 2012 | Jensen et al., 2017; Mortimer et al., 2013, Hoeijenbos et al., 2005 |  |  |
|  | Use an implementation advisor (1) | Healthcare Professionals (270)  Patients (1358) | *Referral to imaging* |  | French et al., 2022 |  |  |
|  |  |  | *Function* |  | French et al., 2022 |  |  |
| **Train and educate stakeholders (38)** | Conduct ongoing training (18) | Healthcare Professionals (958)  Patients (16,054) | *Adherence to implemented intervention* | Rebbeck et al., 2006; Scheel et al., 2002 | Stevenson et al., 2006 |  |  |
|  |  |  | *Uptake of knowledge* | Rebbeck et al., 2006; van Dulmen et al., 2014 | Bussières et al., 2010; Chipchase et al., 2016; Stevenson et al., 2006 | French et al., 2013; Maas et al., 2015 | Stevenson et al., 2004; |
|  |  |  | *Referral to imaging* | Schröder et al., 2022 | French et al., 2013; Suman et al., 2018 |  |  |
|  |  |  | *Referral to secondary care* | Riis et al., 2016 | Schröder et al., 2022; Suman et al., 2018 |  |  |
|  |  |  | *HCP satisfaction* |  | Rebbeck et al., 2006 |  |  |
|  |  |  | *Function* | Becker et al., 2008, Cleland et al., 2009 | Becker et al., 2008; Chipchase et al., 2016; Rebbeck et al., 2006 |  |  |
|  |  |  | *Pain* |  | Cleland et al., 2009 |  |  |
|  |  |  | *Physical activity* |  | Leonhardt et al., 2008 |  |  |
|  |  |  | *Patient Satisfaction* |  | Rebbeck et al., 2006 |  |  |
|  |  |  | *Cost-effectiveness* | Becker et al., 2012 | Jensen et al., 2017; Mortimer et al., 2013 |  |  |
|  | Provide ongoing consultation (11) | Healthcare Professionals (352)  Patients (10,515) | *Adherence to implemented intervention* | Bekkering et al., 2005 (a); Scheel 2002 |  |  |  |
|  |  |  | *Uptake of knowledge* |  | Chipchase et al., 2016 |  |  |
|  |  |  | *Referral to secondary care* | Riis et al., 2016 |  |  |  |
|  |  |  | *Function* | Becker et al., 2008, Cleland et al., 2009 | Bekkering et al., 2005 (b); Becker et al., 2008; Chipchase et al., 2016 |  |  |
|  |  |  | *Pain* |  | Bekkering et al., 2005 (b), Cleland et al., 2009 |  |  |
|  |  |  | *Workability* |  |  |  | Bekkering et al., 2005 (b) |
|  |  |  | *Physical activity* |  | Leonhardt et al., 2008 |  |  |
|  |  |  | *Quality of life* |  | Hoeijenbos et al., 2005 |  |  |
|  |  |  | *Cost-effectiveness* | Becker et al., 2012 | Jensen et al., 2017, Hoeijenbos et al., 2005 |  |  |
|  | Develop educational materials (10) | Healthcare Professionals (2,518)  Patients (15,323) | *Adherence to implemented intervention* | Goldberg et al., 2001; Rebbeck et al., 2006; Sanders et al., 2017; Scheel et al., 2002 | Engers et al., 2005 | Evans et al., 2010 |  |
|  |  |  | *Uptake of knowledge* | Rebbeck et al., 2006 |  |  |  |
|  |  |  | *Referral to imaging* | Schectman et al.,2003; Simula et al., 2021 | Coombs et al., 2021; French et al., 2022 |  |  |
|  |  |  | *Referral to secondary care* | Schectman et al., 2003 | Engers et al., 2005 |  |  |
|  |  |  | *Prescription of analgesics* |  | Engers et al., 2005 |  |  |
|  |  |  | *HCP satisfaction* |  | Rebbeck et al., 2006 |  |  |
|  |  |  | *Function* |  | French et al., 2022; Rebbeck et al., 2006; Simula et al., 2021 |  |  |
|  |  |  | *Patient satisfaction* |  | Rebbeck et al., 2006 |  |  |
|  | Make training dynamic (21) | Healthcare Professionals (1,628)  Patients (3,726) | *Adherence to implemented intervention* | Bekkering et al., 2005 (a); Murray et al., 2015; Peter et al., 2013; Rebbeck et al., 2006; Sanders et al., 2017 | Engers et al., 2005; Stevenson et al., 2006 |  |  |
|  |  |  | *Uptake of knowledge* | Peter et al., 2015, Rebbeck et al., 2006; van Dulmen et al., 2014 | Bussières et al., 2010; Chipchase et al., 2016; Peter et al., 2013; Stevenson et al., 2006 | Maas et al., 2015; French et al., 2013 | Stevenson et al., 2004 |
|  |  |  | *Referral to imaging* | Schröder et al., 2022 | French et al., 2013 |  |  |
|  |  |  | *Referral to secondary care* |  | Engers et al., 2005; Schröder et al., 2022 |  |  |
|  |  |  | *Prescription of analgesics* |  | Engers et al., 2005 |  |  |
|  |  |  | *HCP satisfaction* |  | Rebbeck et al., 2006 |  |  |
|  |  |  | *Function* | Becker et al., 2008 | Becker et al., 2008; Bekkering et al., 2005 (b); Chipchase et al., 2016: Rebbeck et al., 2006 |  |  |
|  |  |  | *Pain* |  | Bekkering et al., 2005 (b) |  |  |
|  |  |  | *Workability* |  |  |  | Bekkering et al., 2005 (b) |
|  |  |  | *Quality of life* |  | Hoeijenbos et al., 2005 |  |  |
|  |  |  | *Physical activity* |  | Leonhardt et al., 2008 |  |  |
|  |  |  | *Patient satisfaction* |  | Rebbeck et al., 2006 |  |  |
|  |  |  | *Cost-effectiveness* | Becker et al., 2012 | Mortimer et al., 2013, Hoeijenbos et al., 2005 |  |  |
|  | Distribute educational materials (30) | Healthcare Professionals (7,863)  Patients (28,657) | *Adherence to implemented intervention* | Bekkering et al., 2005 (a); Goldberg et al., 2001; Murray et al., 2015; Rebbeck et al., 2006; Sanders et al., 2017; Scheel et al., 2002 | Dey et al., 2004; Engers et al., 2005 | Bishop et al 2006; Evans et al., 2010 |  |
|  |  |  | *Uptake of knowledge* | Rebbeck et al., 2006; Suman et al., 2014 |  | French et al., 2013; Maas et al., 2015 |  |
|  |  |  | *Referral to imaging* | O'Connor et al., 2022; Schectman et al., 2003; Schröder et al., 2022; Simula et al., 2021 | Coombs et al., 2021; Dey et al., 2004; French et al., 2013; French et al., 2022; Suman et al., 2018 | Eccles et al., 2001 |  |
|  |  |  | *Referral to secondary care* | Riis et al., 2016; Schectman et al., 2003 | Dey et al., 2004; Engers et al., 2005, Schröder et al., 2022; Suman et al., 2018 |  |  |
|  |  |  | *Prescription of analgesics* |  | Dey et al., 2004; Engers et al., 2005 |  |  |
|  |  |  | *HCP satisfaction* |  | Rebbeck et al., 2006 |  |  |
|  |  |  | *Function* | Becker et al., 2008 | Becker et al., 2008; French et al., 2022; Bekkering et al., 2005 (b); Rebbeck et al., 2006; Simula et al., 2021 |  |  |
|  |  |  | *Pain* |  | Bekkering et al., 2005 (b) |  |  |
|  |  |  | *Workability* |  |  |  | Bekkering et al., 2005 (b) |
|  |  |  | *Quality of life* |  | Hoeijenbos et al., 2005 |  |  |
|  |  |  | *Physical activity* |  | Leonhardt et al., 2008 |  |  |
|  |  |  | *Patient satisfaction* |  | Rebbeck et al., 2006 |  |  |
|  |  |  | *Patient adherence* | Moseng et al., 2019 |  |  |  |
|  |  |  | *Cost-effectiveness* | Becker et al., 2012 | Jensen et al., 2017; Mortimer et al., 2013, Hoeijenbos et al., 2005 |  |  |
|  | Conduct educational meetings (29) | Healthcare Professionals (1,848)  Patients (18,449) | *Adherence to implemented intervention* | Bekkering et al., 2005 (a); Goldberg et al., 2001; Murray et al., 2015; Peter et al., 2013; Rebbeck et al., 2006; Sanders et al., 2017; Scheel et al., 2002 | Engers et al., 2005; Stevenson et al., 2006 |  |  |
|  |  |  | *Uptake of knowledge* | Peter et al., 2015; Rebbeck et al., 2006; Van Dulmen et al., 2014 | Bussières et al., 2010; Chipchase et al., 2016; Peter et al., 2013; Stevenson et al., 2006 | French et al., 2013; Maas et al., 2015 | Stevenson et al., 2004; |
|  |  |  | *Referral to imaging* | Schröder et al., 2022; Simula et al., 2021 | French et al., 2013; Suman et al., 2018 |  |  |
|  |  |  | *Referral to secondary care* | Riis et al., 2016 | Engers et al., 2005; Schröder et al., 2022; Suman et al., 2018 |  |  |
|  |  |  | *Prescription of analgesics* |  | Engers et al., 2005 |  |  |
|  |  |  | *HCP satisfaction* |  | Rebbeck et al., 2006 |  |  |
|  |  |  | *Function* | Becker et al., 2008, Cleland et al., 2009 | Becker et al., 2008; Chipchase et al., 2016; Bekkering et al., 2005 (b): Rebbeck et al., 2006; Simula et al., 2021 |  |  |
|  |  |  | *Pain* |  | Bekkering et al., 2005 (b), Cleland et al., 2009 |  |  |
|  |  |  | *Workability* |  |  |  | Bekkering et al., 2005b |
|  |  |  | *Quality of life* |  | Hoeijenbos et al., 2005 |  |  |
|  |  |  | *Physical activity* |  | Leonhardt et al., 2008 |  |  |
|  |  |  | *Patient satisfaction* |  | Rebbeck et al., 2006 |  |  |
|  |  |  | *Patient adherence* | Moseng et al., 2019 |  |  |  |
|  |  |  | *Cost-effectiveness* | Becker et al., 2012 | Jensen et al., 2017; Mortimer et al., 2013, Hoeijenbos et al., 2005 |  |  |
|  | Conduct educational outreach visits (16) | Healthcare Professionals (5,597)  Patients (14,730) | *Adherence to implemented intervention* | Bekkering et al., 2005 (a); Rebbeck et al., 2006 | Dey et al., 2004 |  |  |
|  |  |  | *Uptake of knowledge* | Rebbeck et al., 2006 |  |  |  |
|  |  |  | *Referral to imaging* | Schectman et al., 2003 | Coombs et al., 2021; Dey et al., 2004; French et al., 2022 |  |  |
|  |  |  | *Referral to secondary care* | Riis et al., 2016; Schectman et al., 2003 | Dey et al., 2004 |  |  |
|  |  |  | *Prescription of analgesics* |  | Dey et al., 2004 | Bruyndonckx et al., 2018 |  |
|  |  |  | *HCP satisfaction* |  | Rebbeck et al., 2006 |  |  |
|  |  |  | *Function* | Becker et al., 2008, Cleland et al., 2009 | Becker et al., 2008; Bekkering et al., 2005 (b); French et al., 2022; Rebbeck et al., 2006 |  |  |
|  |  |  | *Pain* |  | Bekkering et al., 2005 (b), Cleland et al., 2009 |  |  |
|  |  |  | *Workability* |  |  |  | Bekkering et al., 2005b |
|  |  |  | *Physical activity* |  | Leonhardt et al., 2008 |  |  |
|  |  |  | *Patient satisfaction* |  | Rebbeck 2006 |  |  |
|  |  |  | *Patient adherence* | Moseng et al., 2019 |  |  |  |
|  |  |  | *Quality of Life* |  | Hoeijenbos et al., 2005 |  |  |
|  |  |  | *Cost-effectiveness* | Becker et al., 2012 | Jensen et al., 2017, Hoeijenbos et al., 2005 |  |  |
| **Support clinicians (12)** | Remind clinicians (12) | Healthcare Professionals (1,379)  Patients (13,810) | *Adherence to implemented intervention* | Bekkering et al., 2005 (a); Scheel et al., 2002 | Engers et al., 2005 | Bishop et al., 2006 |  |
|  |  |  | *Uptake of knowledge* |  | Bussières et al., 2010 |  |  |
|  |  |  | *Referral to imaging* | Eccles et al., 2001; Schectman et al., 2003 | French et al., 2022 |  |  |
|  |  |  | *Referral to secondary care* | Riis et al., 2016; Schectman et al., 2003 | Engers et al., 2005 |  |  |
|  |  |  | *Prescription of analgesics* |  | Engers et al., 2005 |  |  |
|  |  |  | *Function* |  | Bekkering et al., 2005 (b); French et al., 2022 |  |  |
|  |  |  | *Pain* |  | Bekkering et al., 2005 (b) |  |  |
|  |  |  | *Workability* |  |  |  | Bekkering et al., 2005b |
|  |  |  | *Quality of life* |  | Hoeijenbos et al., 2005 |  |  |
|  |  |  | *Cost-effectiveness* |  | Jensen et al., 2017, Hoeijenbos et al., 2005 |  |  |
| **Engage consumers (4)** | Involve patients/consumers and family members (1) | Healthcare Professionals (462)  Patients (428) | *Adherence to implemented intervention* |  |  | Bishop et al., 2006 |  |
|  | Use mass media (3) | Healthcare Professionals (113)  Patients (6,231) | *Uptake of knowledge* |  |  |  |  |
|  |  |  | *Referral to imaging* |  |  |  |  |
|  |  |  | *Referral to secondary care* | Riis et al., 2016 | Suman et al., 2018 |  |  |
|  |  |  | *Cost-effectiveness* |  | Jensen et al., 2017; Suman et al., 2018 |  |  |
| **Change infrastructure (3)** | Change record systems (3) | Healthcare Professionals (60)  Patients (1,101) | *Adherence to implemented intervention* | Goldberg et al., 2001 |  |  |  |
|  |  |  | *Referral to secondary care* | Riis et al., 2016 |  |  |  |
|  |  |  | *Cost-effectiveness* |  | Jensen et al., 2017 |  |  |
| **NOTE:** As some studies had multiple intervention groups, it is possible that a study can be classified as both significant and not significant simultaneously. | | | | | | | |
