## Supplementary material for "Effectiveness of Implementation Interventions in Musculoskeletal Healthcare: A Systematic Review": Risk of Bias

### Additional file 6 – Risk of Bias Assessment using RoB 2

| ***Table A6.1***  *Summary of Risk of Bias assessment according to the RoB 2 tool* | | | | | | | | | |
| --- | --- | --- | --- | --- | --- | --- | --- | --- | --- |
| **Randomised Controlled Trials** | | **Risk of Bias Domains** | | | | | | | |
| **Study, Year** | **Study design** | **D1** | | **D2** | **D3** | **D4** | **D5** | | **Overall** |
| Bishop et al., 2006 | RCT | 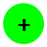 | | 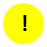 | 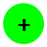 | 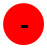 | 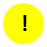 | | 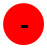 |
| Bussières et al 2010 | RCT | 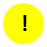 | | 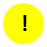 | 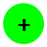 | 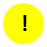 | 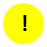 | | 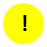 |
| Chipchase et al., 2016 | RCT | 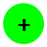 | | 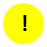 | 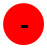 | 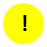 | 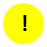 | | 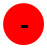 |
| Cleland et al., 2009 | RCT | 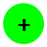 | | 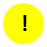 | 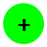 | 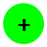 | 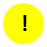 | | 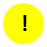 |
| Peter et al., 2013 | RCT | 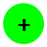 | | 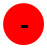 | 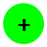 | 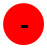 | 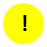 | | 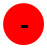 |
| Peter et al., 2015 | RCT | 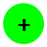 | | 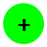 | 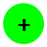 | 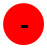 | 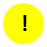 | | 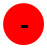 |
| **Cluster Randomised Controlled Trials** | | **Risk of Bias Domains** | | | | | | | |
| **Study, Year** | **Study design** | **D1a** | **D1b** | **D2** | **D3** | **D4** | **D5** | | **Overall** |
| Becker et al., 2012 | Cluster RCT | 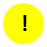 | 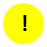 | 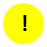 | 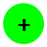 | 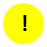 | 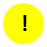 | | 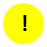 |
| Becker et al., 2008 | Cluster RCT | 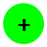 | 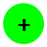 | 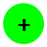 | 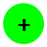 | 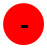 | 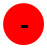 | | 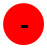 |
| Bekkering et al., 2005 (a) | Cluster RCT | 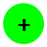 | 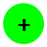 | 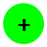 | 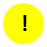 | 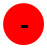 | 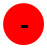 | | 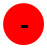 |
| Bekkering et al., 2005 (b) | Cluster RCT | 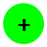 | 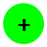 | 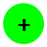 | 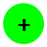 | 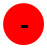 | 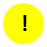 | | 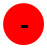 |
| Bruyndonckx et al., 2018 | Cluster RCT | 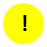 | 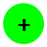 | 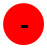 | 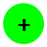 | 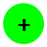 | 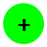 | | 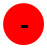 |
| Coombs et al., 2021 | Cluster RCT | 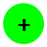 | 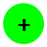 | 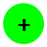 | 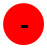 | 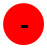 | 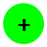 | | 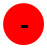 |
| Dey et al., 2004 | Cluster RCT | 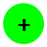 | 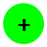 | 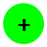 | 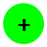 | 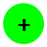 | 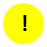 | | 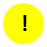 |
| Eccles et al., 2001 | Cluster RCT | 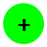 | 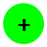 | 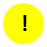 | 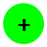 | 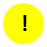 | 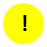 | | 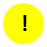 |
| Engers et al., 2005 | Cluster RCT | 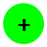 | 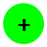 | 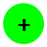 | 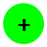 | 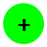 | 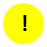 | | 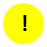 |
| Evans et al., 2010 | Cluster RCT | 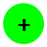 |  |  |  |  |  | |  |
| French et al., 2013 | Cluster RCT |  |  |  |  |  |  | |  |
| French et al., 2022 | Cluster RCT |  |  |  |  |  |  | |  |
| Goldberg et al., 2001 | Cluster RCT |  |  |  |  |  |  | |  |
| Hoeijenbos et al., 2005 | Cluster RCT |  |  |  |  |  |  | |  |
| Jensen et al., 2017 | Cluster RCT |  |  |  |  |  |  | |  |
| Leonhardt et al., 2008 | Cluster RCT |  |  |  |  |  |  | |  |
| Maas et al., 2015 | Cluster RCT |  |  |  |  |  |  | |  |
| Mortimer et al., 2013 | Cluster RCT |  |  |  |  |  |  | |  |
| Moseng et al., 2019 | Cluster RCT |  |  |  |  |  |  | |  |
| Murray et al., 2015 | Cluster RCT |  |  |  |  |  |  | |  |
| O'Connor et al., 2022 | Cluster RCT |  |  |  |  |  |  | |  |
| Rebbeck et al., 2006 | Cluster RCT |  |  |  |  |  |  | |  |
| Riis et al., 2016 | Cluster RCT |  |  |  |  |  |  | |  |
| Sanders et al., 2017 | Cluster RCT |  |  |  |  |  |  | |  |
| Schectman et al., 2003 | Cluster RCT |  |  |  |  |  |  | |  |
| Scheel et al. 2002 | Cluster RCT |  |  |  |  |  |  | |  |
| Schröder et al., 2022 | Cluster RCT |  |  |  |  |  |  | |  |
| Simula et al., 2021 | Cluster RCT |  |  |  |  |  |  | |  |
| Stevenson et al., 2004 | Cluster RCT |  |  |  |  |  |  | |  |
| Stevenson et al., 2006 | Cluster RCT |  |  |  |  |  |  | |  |
| Suman et al., 2018 | Cluster RCT |  |  |  |  |  |  | |  |
| van Dulmen et al., 2014 | Cluster RCT |  |  |  |  |  |  | |  |
| **Outcome domains:**  **D1:** Randomization process  **D1a:** Randomization process  **D1b:** Risk of bias arising from the timing of identification or recruitment of participants  **D2:** Deviations from intended interventions  **D3:** Missing outcome data  **D4:** Measurement of the outcome  **D5:** Selection of the reported result | | | | | | | | **Judgement:** | |
|  |  |  |  |  |  |  |  | High | |
|  |  |  |  |  |  |  |  | Some concerns | |
|  |  |  |  |  |  |  |  | Low | |
